# Facilitators and Barriers to Community-Based Livestock Abortion Reporting for Rift Valley Fever Surveillance in Uganda: A COM-B Analysis

**DOI:** 10.1101/2025.08.01.25332592

**Authors:** Abel W. Walekhwa, Andrew JK Conlan, Lydia N. Namakula, Brenda Nakazibwe, Salome A. Bukachi, James L.N. Wood, Lawrence Mugisha

**Affiliations:** Diseases Dynamics Unit, Department of Veterinary Medicine, University of Cambridge, UK; IDEMU Mathematical Modelling Unit, Makerere University School of Public Health, Kampala; Makerere University School of Public Health, P.O Box 7072, Kampala, Uganda; Pathogen Economy Bureau, Science, Technology and Innovation Secretariat, Office of the President, Kampala, Uganda; Makerere University College of Veterinary Medicine, Animal Resources and Biosecurity, Kampala, Uganda; Ecohealth Research Group, Conservation and Ecosystem Health Alliance (CEHA), Kampala, Uganda; Department of Anthropology, Gender and African Studies, University of Nairobi, Kenya

**Keywords:** Capabilities, Opportunities, Motivation, Reporting, Livestock abortions, Uganda

## Abstract

Rift Valley Fever Disease (RVFD) causes substantial economic losses via livestock abortion storms, yet community-based abortion reporting for early warning remains suboptimal in endemic settings. This study analysed the behavioural determinants of livestock abortion reporting to inform RVF surveillance interventions in Uganda.

We conducted a cross-sectional study using qualitative data collection methods in Isingiro District, Uganda, employing the Capability, Opportunity, Motivation–Behaviour (COM-B) framework. We conducted 29 key informant interviews with national/district policymakers and technical officers and 17 focus group discussions involving livestock owners, abattoir operators, and local leaders. Transcripts underwent deductive thematic analysis using NVivo-12, with independent coding verification and member checking. Barriers and facilitators were ranked by frequency and thematic emphasis, and a comparative analysis was conducted across participant groups.

The three most critical barriers were: poor community knowledge about RVF, with all confirmed cases initially submitted as Ebola suspects; veterinary personnel understaffing, with officers covering four or more sub-counties; and absence of post-reporting feedback, the most powerful demotivator. The three main facilitators were: mobile phone platforms enabling real-time reporting, existing community governance structures facilitating information sharing, and fear of economic and cultural livestock losses motivating reporting despite knowledge deficits. Comparative analysis revealed divergent perspectives: policymakers framed economic concerns macroeconomically, while communities experienced them as household-level losses. COM-B interconnectedness analysis demonstrated that the absence of feedback created disengagement, fear partially compensated for knowledge gaps, and single-domain interventions failed without addressing all three COM-B components.

Our study recommends that sustainable abortion reporting requires simultaneous intervention across capability, opportunity, and motivation domains. Priority interventions could include: integrating reporting into Uganda’s electronic surveillance platforms, co-developing local RVF names with communities, establishing reliable feedback mechanisms, addressing veterinary staffing gaps in remote areas, and implementing no-blame reporting protocols. Effective RVF surveillance depends on reinforcing existing community structures while ensuring responsive feedback that validates reporters’ contributions.

## 1.0 Introduction

Rift Valley Fever disease (RVFD) is caused by the Rift Valley Fever (RVF) virus, transmitted mainly via mosquito vectors (*Aedes* and *Culex spp)* (Hartman, 2017). The most notable symptom in livestock with RVF is abortion “storms” (Ali et al., 2012). The occurrence of abortion storms could be used to institute syndromic surveillance for RVF (Thomas et al., 2022; Walekhwa et al., 2025).

Surveillance for livestock abortions, as proposed by Thurmond and Picanso (Thurmond and Picanso, 1990), hinges on effective reporting. Community-based reporting refers to a participatory surveillance approach that empowers local livestock stakeholders to recognise and report livestock abortion events in a timely and structured manner, thereby strengthening early detection, response and control of diseases like RVF (Giorgio et al., 2023). This reporting, if consistently done, enables real-time or near real-time detection (Gachohi et al., 2024a; Thomas et al., 2022; Walekhwa et al., 2025).

In our study, we sought to understand the facilitators and barriers to community-based reporting of livestock abortions for RVF surveillance in Isingiro District, Uganda. A number of short-term research studies in East Africa, including our own demonstration of feasibility in Uganda (Walekhwa et al., 2025), have highlight the potential of this approach. A notable research study in the arid and semi-arid lands of Kenya used self-reporting by phone to quantify the burden of livestock abortions. The leadership of Kenya Veterinary Services (KVS), the availability of phones, and the training of local animal health practitioners were the enabling factors for its success (Gachohi et al., 2024b). However, there is scant information on what happened after this research study concluded. This highlights a critical gap that Worsley-Tonks et al. (2025) also highlight: the need for sustainable, long-term integration of community-based surveillance into national health systems (Worsley-Tonks et al., 2025).

Another research study by Thomas et al. enabled the detection of ten different pathogens, with RVF identified as the leading cause of abortion (Lankester et al., 2024; Semango et al., 2024; Thomas et al., 2022). However, a critical gap remains in understanding the specific facilitators and barriers that influence sustained reporting behaviour. To address this, we adopted an implementation science approach using the COM-B model (Michie et al., 2011) to systematically analyse the behavioural determinants of abortion reporting. This work contributes to a growing body of knowledge advancing community-based surveillance systems (Worsley-Tonks et al., 2025).

## 2.0 Methods

### 2.1 Ethics Statement

Ethical approval for this study was sought and received from Makerere University School of Public Health (Reference No: SPH-2022-364). Administrative clearance from the Isingiro District local government’s administration was also required and provided by the Chief Administrative Officer’s office (dated 20^th^ March 2023). All methods were performed in accordance with relevant national and local guidelines and regulations.

Respondents were taken through a consenting process and were provided with information about the study. Their voluntary participation and consent was recorded in writing. To enhance understanding, we translated all the data collection tools and informed consent forms into the local language (Runyankole), which was the most spoken language in the Isingiro District. All the data were de-identified to protect the identity of our informants and stored in an encrypted and password-protected external drive of the corresponding author. The emerging quotes were anonymised by using aggregated descriptors.

### 2.2 Study design

We employed a qualitative research design with deductive thematic analysis.

#### Data Collection Period

Data were collected between 22^nd^ March 2023 and 4^th^ July 2023.

### 2.3 Description of the study sites

The location of this study, Isingiro District, Uganda, is an area where previous RVF outbreaks have been reported (Bakamutumaho et al., 2025; Muema et al., 2021; Ndishimye et al., 2024; Nyakarahuka et al., 2023). The specific 17 sub-counties and town councils of Masha, Bugango TC, Kikagati, Kashumba, Kakamba, Endiizi, Endiizi TC, Mbaare, Ngarama, Ruborogota, Rushasha, Rugaga, Rugaga TC, Rwanjogyera, Rwetango, Nyakitunda and Nakivale refugee settlement were selected as they carry out livestock rearing practices associated with a high risk of RVF (Agaba et al., 2025). Most of the respondents were community representatives from the study area and Isingiro District local government workers. Isingiro District is in southwestern Uganda (0.8435° S, 30.8039° E), about 297 kilometres from the capital city, Kampala (Fig. 1).

**Fig 1:**
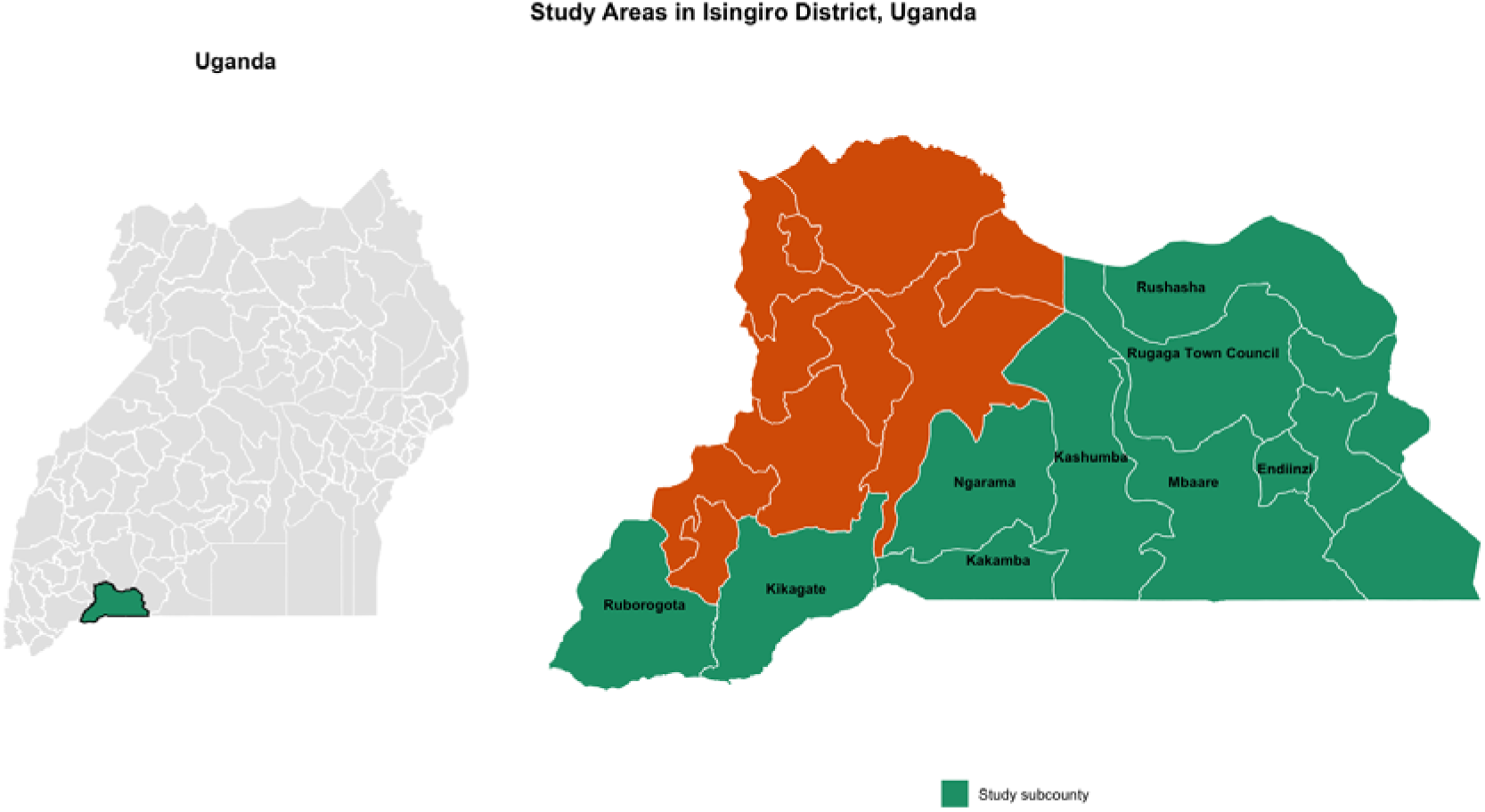
Map of Uganda showing Study sub-counties, Isingiro District, Uganda

Isingiro District is characterised by livestock rearing as a major livelihood for the majority of residents (Adonia, 2013; Bwengye et al., 2023; Mubiru et al., 2023a). The district also neighbours Lake Mburo National Park, which harbours wildlife species susceptible to RVF (Agaba et al., 2025). Both wildlife and livestock share common water sources along the shores of Lake Mburo and the Kagera River, which spans the district. Isingiro District has an estimated cattle, goat, and sheep population of 368,246, 422,108, and 88,621, respectively (UBOS, 2024), and a human population of 635,077 based on the human census (UBOS, 2024).

The district experiences a tropical savanna climate with an average annual rainfall of 1,200 mm, and a temperature range of 17–30° C (Kweyu et al., 2023; Nagasha et al., 2019). The district has two rainy seasons each year: March to April and September to November (Taremwa et al., 2020). These climatic conditions play a critical role in the spread of RVF (Mpeshe et al., 2014).

### 2.4 Theoretical framework of this study

We employed an implementation science (IS) approach grounded in the COM-B model to investigate the facilitators and barriers to community-based abortion reporting for RVF surveillance. IS provides a structured approach for translating evidence-based interventions into real-world settings and is especially relevant for strengthening health systems in low - and middle-income countries (LMICs) (Bauer and Kirchner, 2020). The use of a robust theoretical framework like the COM-B model is a significant strength of this study, as it allows for a systematic and comprehensive analysis of the factors influencing behaviour change.

The COM-B model, developed by Michie et al. (Michie et al., 2011), posits that behaviour change occurs when individuals or groups have the Capability, Opportunity, and Motivation to perform a behaviour. In the context of this study, the target behaviour was the reporting of livestock abortions by community members.

Capability refers to the psychological and physical capacity to engage in a behaviour. In the context of RVF surveillance in Uganda, this includes knowledge and training of veterinary and health officers on RVF case recognition, abortion reporting systems, and sample handling procedures. It also involves awareness of surveillance and reporting channels such as the electronic Integrated Disease Surveillance and Response (eIDSR) system and the World Organisation for Animal Health – World Animal Health Information System (OIE-WAHIS). These factors determine whether frontline personnel have the skills and understanding necessary to detect and report RVF cases accurately.

Opportunity encompasses external factors that make the behaviour possible or prompt it. For RVF surveillance, these include the availability of logistics and resources (e.g. transport, sample collection kits, and laboratory networks), effective communication infrastructure (phones and radios) and coordination between human and animal health sectors under the One Health framework. Community-based surveillance systems and supportive governance structures also create opportunities for timely detection and information sharing.

Motivation involves the mental processes that energise and direct behaviour, including both intrinsic and extrinsic incentives. In the Ugandan context, this is reflected in the perceived importance of RVF surveillance, fear of economic losses due to livestock deaths, cultural obligations to protect community livelihoods, and perceptions of disease severity. Motivation can also be influenced by recognition or incentives for timely reporting, as well as by emotional factors such as fear of blame or burnout associated with high workloads.

Behaviour refer to specific actions an individual or group performs (or does not perform) that interventions aim to change. For our case, this was abortion reporting by community-level stakeholders.

The strength of the COM-B model lies in its ability to illustrate how these components are interdependent and dynamic (Cecilia et al., 2022; Connolly et al., 2021). This implies that interventions targeting one domain (such as training to improve capability) must also ensure that opportunities for action and motivational reinforcements (such as feedback and visible responses) are present to sustain behavioural change. This integrative understanding is particularly crucial in the surveillance of zoonotic diseases, where both human and animal health systems interact. This resonates with the core message of Worsley-Tonks et al. (2025), who argue that successful community-based surveillance requires a holistic approach that addresses not only the capacity of community members but also the broader systemic and structural factors that enable or hinder their participation (Worsley-Tonks et al., 2025).

#### 2.4.1 Intervention (Training)

Before qualitative data collection for this study, a community sensitisation training programme was conducted in Isingiro District as part of the broader abortion surveillance pilot study (Walekhwa et al., 2025). This training is not the “intervention” being evaluated in this study; rather, it established a contextual foundation that informed participants’ awareness and experiences, which were subsequently explored through the qualitative inquiry.

The office of the District Veterinary Officer (DVO), responsible for animal health surveillance at the sub-national level, co-developed the training material with the corresponding author. The developed instructional material included the local authority DVO’s telephone number, which farmers could contact in case of abortions at their farms. Local leaders distributed these targeted messages during community funerals and barazas (public community meetings where residents gather to discuss local issues), intending to encourage the reporting of abortions. The instructional materials used in this training are provided (S1 Appendix).

The purpose of describing this training here is threefold: (1) to provide transparency about the implementation context in which the qualitative data were collected, (2) to acknowledge that some participants in the KIIs and FGDs could have been exposed to this sensitisation through neighbouring sub-counties/locations, which may have influenced their knowledge and attitudes, and (3) to situate the facilitators and barriers identified in this study within a real-world setting where initial capacity-building efforts had already been undertaken in a neighbouring sub-counties. This is especially important because the farmers/stakeholders interact in social gathering events, and they tend to discuss livestock-related interventions by the government or different stakeholders like ours. Although efforts were made to first interview the stakeholders before being trained, we cannot fully rule out the possibility of contamination from neighbouring sub-counties.

A one-day training intervention was developed and delivered to stakeholders across multiple sub-counties, followed by three weeks of reinforced messaging. The training was designed to build practical skills and knowledge for syndromic surveillance. The core curriculum covered: (a) the clinical recognition of livestock abortions and their importance as a sentinel event for zoonotic diseases such as RVF. (b) Signs and symptoms of RVF in both animals and humans, according to national surveillance guidelines. (c) The specific action to take upon identifying an abortion: calling a dedicated telephone number for the local authority DVO’s office, which was printed on the instructional materials. (d) The purpose and benefits of a community surveillance system. We conducted in-person training sessions in English, with simultaneous translation into the local language (Runyankole) provided by a native technical officer to ensure comprehension. This training was conducted among 900 stakeholders, including seven district officials, 72 sub-county technical officers, 38 community health workers, 788 livestock owners, abattoir staff, and local council leaders (S1 Table). Qualitative data collection for this study commenced approximately 3-4 days after the conclusion of this sensitisation campaign in the neighbouring sub-counties.

### 2.5 Data collection

#### 2.5.1 Participants and Sampling

First, census sampling was conducted for all livestock-rearing sub-counties (17/33). We then used gatekeepers (community leaders) to select participants from communities in regions affected by RVF and ensured diversity in gender, occupation, age, economic status and geographical distribution. The selection process emphasised individuals whose livelihoods, beliefs, or practices intersect with RVF risk and the consequences of abortions. These included livestock owners, community leaders, meat handlers, private veterinary practitioners, subnational leaders (sub-county leaders) and farm managers/owners. Those who accepted the invitation were guided to attend a one-day training. From the participants who turned up, a subset was identified and invited to participate in the study. The majority were aged 30-39 years (29%), with primary (34%) and secondary (32%) education being the most common level of schooling. This sampling strategy, which combines census and purposive sampling, is appropriate for the research question and the specific context of the study.

The use of gatekeepers to identify participants is a common and accepted practice in qualitative research, particularly in community-based studies (Crowhurst and Kennedy-Macfoy, 2013; Rankin and McFadyen, 2016). To this end, data were collected using Key Informant Interviews (KIIs) and Focus Group Discussions (FGDs) guides (S2 Appendix and S3 Appendix).

#### 2.5.2 Procedure of Focus Group Discussions

A total of 17 FGDs (from 17 different sub-counties) were conducted. We chose the FGDs to enable understanding of group-level factors for the intervention and to create an interactive environment that would allow for further probing (Setia, 2017). We considered respondents of similar socio-economic status in the community so that they could freely engage in the discussions. The choice of respondents was purposeful, with stakeholders selected given their active roles in human and animal surveillance efforts in Uganda. The interviews and discussions were held in various sites that were convenient to the respondents. All FGDs were facilitated by a leader, who was selected at the start of each session. The sessions were held in the local language (Runyankole) and moderated by a research assistant who was a native speaker. Each FGD session lasted approximately 60 minutes (range: 52 – 74 minutes), providing adequate time for all participants to share their perspectives while maintaining engagement throughout the discussion. The use of FGDs is a key strength of this study, as it allows for the exploration of shared experiences and social norms related to abortion reporting (O.Nyumba et al., 2018). This aligns with the emphasis in Worsley-Tonks et al. (2025) on the importance of understanding the cultural and social context of community-based surveillance (Worsley-Tonks et al., 2025).

#### 2.5.3 Procedure of Key Informant Interviews

For KIIs, 29 policymakers and senior technical officials from district (sub-national) and national levels, and from both public and private agencies (ministries, academia, Food and Agriculture Organisation) were enrolled. Some of the respondents included the incident commander for disease outbreaks; the director of the National Institute of Public Health – Ministry of Health; ministry technical staff, animal health surveillance officers, laboratory technicians, environmental health officers, senior veterinary officers and field epidemiologists. The use of KIIs aimed to explore high-level expert opinions on this subject, providing strategic and contextual explanations for facilitators and barriers to community-based reporting, as recommended by Tumusiime et al. (Tumusiime et al., 2023a). For the KIIs, apart from four informants who requested a virtual interview, the rest of the interviews were conducted physically in locations convenient to respondents. As part of the participant recruitment process, each participant was taken through a consenting process (using a prepared consent form), and only those who gave written informed consent were included in the study. KIIs lasted an average of 47 minutes (range: 32-68 minutes). The four virtual interviews were comparable in duration to in-person interviews (mean: 44 minutes vs 48 minutes, respectively). No participant was interviewed more than once. Each key informant participated in a single interview, and each focus group participant attended only one FGD session. Repeat interviews were not conducted as the study aimed to capture perspectives at a single time point following the training intervention, and saturation was achieved within the initial round of data collection.

We conducted interviews until saturation was attained at the 29^th^ KII interview and 17^th^ FGD session, as respondents provided no new information. All individuals invited to participate in the study accepted the invitation, resulting in a 100% participation rate with no refusals or dropouts. For the KIIs, all 29 invited policymakers and technical officers consented to participate. For the FGDs, all 17 scheduled focus group discussions proceeded with full attendance from invited participants. This high acceptance rate can be attributed to the prior training intervention, which established rapport and demonstrated the study’s value to the community, as well as the use of respected gatekeepers (community leaders) who facilitated trust between researchers and potential participants. No participants withdrew from the study after providing consent.

#### 2.5.4 Interviewer Characteristics and Team Composition

The data collection team comprised five interviewers: two males and three females. The gender-balanced team was intentionally composed to facilitate comfortable discussions with diverse participant groups, as community norms in the study area sometimes influence disclosure based on interviewer-participant gender dynamics. The team included:

1. The corresponding author (AWW) – male senior epidemiologist who led KIIs with policymakers and technical officers.
2. A female research assistant (Mellon Ainembabazi) – native Runyankole speaker and who also served as a female youth local council leader for Bugango Town Council, listened and guided the moderators of all FGDs.
3. Two additional female research assistants supported notetaking and logistics during FGDs.
4. One male research assistant supported district-level KIIs and technical coordination. The gender composition proved advantageous: female participants in FGDs appeared more forthcoming when discussing sensitive topics (such as cultural practices around livestock and dowry) with the female moderator, while male policymakers engaged readily with the male corresponding author during KIIs. All interviewers received standardised training on qualitative data collection techniques, maintaining consistency in approach while leveraging gender-appropriate facilitation.

### 2.6 Data Processing and Quality Assurance

During the entire data collection process, all interviews were recorded. The audio recordings were later transcribed verbatim by two trained, experienced research assistants. For quality assurance in audio transcriptions, the corresponding author sampled six audios (three from each research assistant) and listened to the audio in comparison to the transcript, which was found to be consistent. Beyond this sampling, we conducted multiple rounds of validations to ensure credibility. We cross-referenced each theme with verbatim quotations to ensure it accurately represented the respondents’ perspectives. Multiple researchers reviewed themes independently to eliminate potential biases and inconsistencies. This refinement ensured that the final themes were precise and accurately reflected the data. The transcripts were then analysed using deductive thematic analysis with NVivo-12 software (Edhlund and McDougall, 2017) guided by the COM-B model.

To enhance the credibility and trustworthiness of findings, we conducted member checking with a subset of participants following initial analysis. This process involved:

**Presentation of preliminary findings at three intervals.** First, on the last day of data collection, findings were presented to 23 Isingiro district-level officials as part of the exit from fieldwork. Secondly, six months later, findings were presented to 18 district-level stakeholders and sub-county veterinary staff, including six livestock owners. Finally, findings were presented to 13 staff of the National Animal Diseases Diagnostics and Epidemiology Centre (NADDEC) at the Ministry of Agriculture, Animal Industries and Fisheries.

The first two sessions were conducted both in Runyankole and English, facilitated by the district veterinary officer. The national level sessions with the Ministry of Agriculture were facilitated by the male facilitator (the corresponding author).

#### Validation Process

During these meetings, we presented the key themes organised by the COM-B framework (facilitators and barriers in each domain) using simple language and illustrative quotes (anonymised). Participants were encouraged to:

- Confirm whether the themes accurately reflected their experiences.
- Challenge any interpretations they felt were incorrect or incomplete.
- Provide additional context or examples to enrich the findings.
- Prioritise which barriers they considered most critical.

#### Outcome of member checking

Participants unanimously confirmed that the identified themes captured their experiences accurately. Importantly, they strongly emphasised that the absence of feedback mechanisms was the single most demotivating factor – a perspective that validated our analytical emphasis on this theme. Several livestock owners added nuanced examples of how delayed responses affected their reporting decisions, which were incorporated into the final analysis. No participant contested any themes, and there was consensus that the facilitators and barriers presented reflected community realities.

#### Independent Validation by Research Assistants

Two research assistants who were involved in data collection but not in the coding team (the female FGD facilitator and male note taker) independently reviewed the final thematic framework. They confirmed that the themes aligned with their field observations and recalled specific discussions that supported each major finding. This external verification provided an additional layer of credibility, as these individuals had direct exposure to participant narratives during data collection but were uninvolved in the analytical decisions that could introduce bias.

#### Reflexivity and Researcher Positionality

This stage involved a researcher-led thematic analysis informed by the Braun and Clarke (Braun and Clarke, 2006) framework. We conducted a primarily deductive thematic analysis guided by the COM-B framework, while remaining open to inductive sub-themes that emerged from the data. The process followed a structured, iterative approach:

a. Familiarisation and Initial Coding: We immersed ourselves in the data by repeatedly reading all the transcripts while listening to the audio recordings. The initial coding was conducted using both deductive and inductive approaches. The COM-B model provided the primary deductive framework, with Capability, Opportunity, and Motivation serving as parent codes. Within this framework, we remained open to emergent codes that captured nuanced aspects of each domain.
b. Systematic Code Development: Using NVivo-12 software (Edhlund and McDougall, 2018), we developed a structured coding framework. The transcripts were systematically coded by identifying meaning units (text segments expressing a complete thought or concept) and assigning the appropriate codes. For example, text segments discussing “knowledge of abortion causes” were coded under Capability → Psychological Capacity, while segments about “mobile phone access” were coded under Opportunity → Physical Environment. The systematic coding process generated a comprehensive codebook containing 99 distinct codes with 514 total references across all transcripts, demonstrating the depth and breadth of the qualitative inquiry (S4 Appendix for the complete codebook).
c. Theme Development and Refinement: The coded data were collated and analysed for patterns within and between COM-B domains. The preliminary themes were developed by grouping related codes and examining their relationships. This process involved iterative refinement through team discussions with Lydia N. Namakula and Brenda Nakazibwe, who independently reviewed a subset of transcripts (n = 8, 27.6 % of the total) to challenge initial interpretations and enhance analytical rigour.
d. Theme Review and Validation: The preliminary thematic framework was reviewed for internal homogeneity (whether data within themes cohered meaningfully) and external heterogeneity (whether themes were distinct from each other). This involved cross-checking themes against the original transcripts to ensure that they accurately represented participants’ meanings. Discrepancies in interpretation were resolved through consensus discussions.
e. Theme Definition and Narration: The final themes were clearly defined and named to capture their essence. A codebook (S4 Appendix) was developed, documenting each theme’s definition, characteristics, representative quotes, and frequency of references. This codebook guided the final analysis and writing of the result narrative (Table 1).

**Table 1:** Distribution of codes and references across COM-B Domains.

| COM-B Domain | Number of Codes | Total References |
| --- | --- | --- |
| Capability (Facilitators) | 15 | 98 |
| Capability (Barriers) | 18 | 156 |
| Opportunity (Facilitators) | 22 | 124 |
| Opportunity (Barriers) | 16 | 87 |
| Motivation (Facilitators) | 14 | 76 |
| Motivation (Barriers) | 14 | 73 |
| **Total** | **99** | **514** |

To ensure analytical rigour, multiple quality assurance measures were implemented. For transcription quality, the corresponding author used a systematic validation approach, randomly selecting and thoroughly reviewing six transcripts (13% of the total) by comparing audio recordings against transcripts. This exceeds the minimum 10% validation rate recommended for qualitative research (Reynolds et al., 2011). Independent coding verification by two analysts (co-authors) achieved 87% inter-rater agreement on primary codes, with discrepancies resolved through consensus discussion, thereby enhancing the trustworthiness of the thematic analysis.

To address potential bias arising from researcher characteristics and prior experiences, we maintained reflexive journals throughout data collection and analysis and engaged in systematic positionality reflection. The following researcher characteristics are relevant to interpreting this study:

**a) Corresponding Author (AWW):** Abel W. Walekhwa is an epidemiologist with 12 years of field experience in Uganda’s livestock and health systems. His professional roles include serving as a member of the High-Level Expert panel on One Health with four UN agencies (FAO, UNEP, WHO, WOAH), which shapes the perspective toward recognising systemic barriers and policy-level solutions. He previously worked on disease outbreak responses for eight years, including in Sheema District – a neighbouring district to the study area – providing him with a grounded understanding of the health systems, cultural norms, and operational challenges in southwestern Uganda. In addition, he has over 17 years of experience with livestock rearing (grazing cattle during upbringing). This insider knowledge gave him credibility with participants and enabled an understanding of the practical realities of livestock management, economic pressures facing farmers, and the cultural significance of cattle (including dowry practices). However, this background also risked bias: It was possible to overemphasise technical barriers (diagnostic capacity, transport) based on his epidemiology training or make assumptions about farmer experiences based on his own background rather than letting participants’ narratives guide interpretation.

#### Mitigation Strategies

*Independent coding verification:* Two co-authors (Lydia N. Namakula and Brenda Nakazibwe), who were not involved in data collection, independently reviewed themes and coding decisions. Brenda’s social sciences perspectives challenged Abel’s technical biases – for example, she noted early in analysis that he was undervaluing motivational and cultural factors, which prompted re-examination of the data and ultimately led to foregrounding feedback mechanisms and economic sensitivity as primary barriers.

*Social Sciences mentorship:* Prof. Salome A. Bukachi (anthropologist) provided technical mentorship throughout analysis, consistently drawing attention to cultural dimensions that Abel had minimised.

*Reflexive journaling:* The corresponding author documented after each interview and during the analysis phases how his positionality might shape interpretations. For instance, he noted after early KIIs with policymakers that he was empathising strongly with their resource constraint narratives; journaling helped the corresponding author to consciously balance these perspectives with community-level views from FGDs.

*Participant validation:* Member checking ensured that the interpretation resonated with participants themselves, providing an external check on researcher bias.

**b) Female FGD Facilitator (Mellon Ainembabazi):** As a native Runyankole speaker and community leader, she brought cultural fluency that facilitated open discussions. Her gender was advantageous for discussing sensitive topics with female participants. She maintained her own reflexive notes, documenting instances where participants’ body language or off-record comments provided context for interpreting verbal responses. Her independence from the coding team meant she could provide candid feedback on whether written interpretations matched her recall of discussion tone and emphasis.

**Research Assistants (Validation Role):** The two research assistants who reviewed final themes (female facilitator and male note-taker) had extensive field experience in the region but were not involved in coding decisions (Birt et al., 2016; Candela, 2019). This separation of roles meant they served as validators – confirming that themes reflected field realities without being invested in analytical choices.

The combination of insider perspectives (AWW’s livestock background, FGD facilitator’s cultural fluency) and outsider analytical distance (independent coders, social sciences mentorship) created productive tension that strengthened analytical rigour. We explicitly discussed during team meetings how our positionalities might shape interpretation, and we actively sought disconfirming evidence, cases where participants contradicted emerging themes. This systematic reflexivity ensured that findings remained grounded in participants’ accounts rather than the researcher’s assumptions.

### 2.8 Reporting of findings

The findings are reported following the Consolidated Criteria for Reporting Qualitative Research (COREQ) guidelines (Tong et al., 2007) to ensure transparency and completeness. To maintain participant anonymity while preserving the analytical value of distinguishing between respondent categories, a standard coding scheme was developed and applied to all quotations presented in this manuscript (Table 2).

**Table 2:**
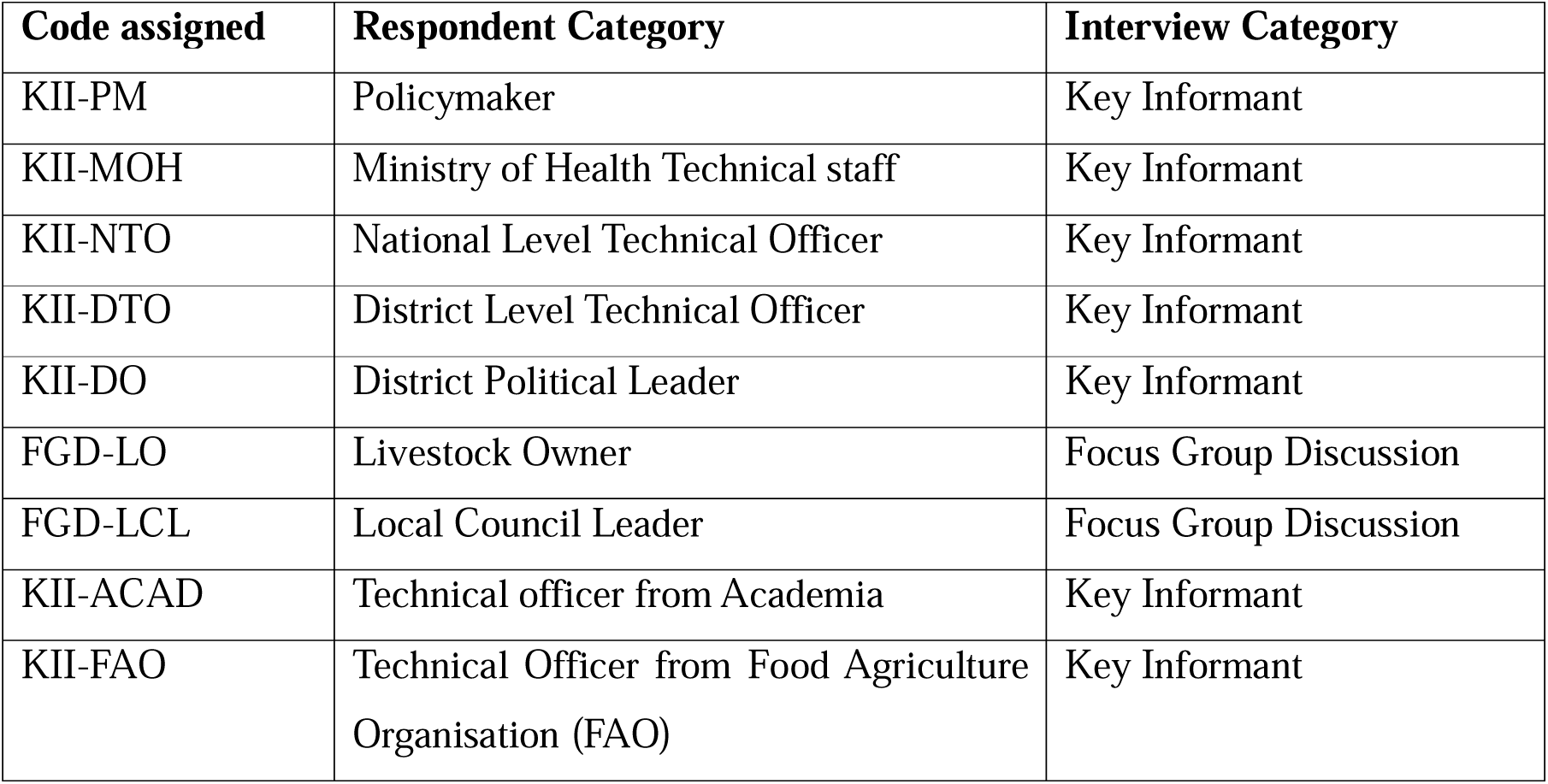
Participant Coding Scheme: KII denotes Key Informant Interviews, FGD denotes Focus Group Discussions.

| Code assigned | Respondent Category | Interview Category |
| --- | --- | --- |
| KII-PM | Policymaker | Key Informant |
| KII-MOH | Ministry of Health Technical staff | Key Informant |
| KII-NTO | National Level Technical Officer | Key Informant |
| KII-DTO | District Level Technical Officer | Key Informant |
| KII-DO | District Political Leader | Key Informant |
| FGD-LO | Livestock Owner | Focus Group Discussion |
| FGD-LCL | Local Council Leader | Focus Group Discussion |
| KII-ACAD | Technical officer from Academia | Key Informant |
| KII-FAO | Technical Officer from Food Agriculture Organisation (FAO) | Key Informant |

Results were organised according to the COM-B framework, with facilitators and barriers presented separately for each domain (Capability, Opportunity, Motivation). Representative quotes are provided to illustrate key themes, with participant codes shown above to indicate the data source while maintaining anonymity.

## 3.0 Results

### 3.1 COM-B Interconnectedness

Our study revealed several nuanced interactions between the three COM-B components. *Opportunity moderates motivation;* the absence of formal responses to submitted reports, through mechanisms, was the most frequently cited demotivator (12 responses), creating a vicious cycle where reporters perceive their effort as futile and disengage. Conversely, structured community meetings (LC 1 gatherings) provided recurring opportunities that simultaneously enhanced capability through knowledge exchange.

*Motivation compensates for limited capability* in striking ways – fear of cultural disruption (missed dowries) and economic losses drove reporting even where knowledge of RVF was poor, while the economic sensitivity of trade restrictions created powerful disincentives that overrode civic duty. *Capability enables sustained motivation* through trust; when livestock owners understood reporting purposes and witnessed functional veterinary laboratories responding, their confidence and willingness to report improved markedly. The profound knowledge gap (exemplified by all confirmed RVF cases initially submitted as Ebola suspects) demonstrates how psychological capability deficits fundamentally undermine the surveillance cascade from its foundation.

In summary, successful community-based surveillance requires simultaneous attention to all three components. Interventions targeting only one domain (e.g., capability through training) is likely to fail without addressing transport infrastructure and veterinary understaffing (opportunity) or establishing transparent feedback loops (motivation). Mobile technologies such as MT-Trac emerge as critical enablers across domains, but their effectiveness depends on robust response systems. Cultural and economic factors play pivotal motivational roles, with livestock serving as economic assets and social capital. Systemic barriers, particularly poor One Health coordination and the absence of local disease nomenclature, require policy-level intervention that recognises the profound interconnectedness of capabilities, opportunities, and motivations driving reporting behaviour.

### 3.2 Comparative perspectives Across Participant Groups

Analysis revealed both convergence and divergence between key informants (policymakers/technical officials) and the focus group participants’ (community members and local leaders) perspectives. Both groups unanimously identified poor community knowledge about RVF as the most fundamental barrier to surveillance (55 references), with KII noting that “all confirmed RVF cases were submitted as Ebola suspects” and FGD participants admitting “we don’t know the disease”. Similarly, both groups highlighted severe understaffing of veterinary personnel, with FGD participants reporting waiting “five months without seeing the doctor” and KIIs confirming one officer covering multiple sub-counties (S5 Appendix).

However, notable differences emerged in how barriers were conceptualised. KIIs emphasised systemic and structural challenges, including inter-ministerial coordination failures, One Health platform weaknesses, and national policy inertia. For example, KIIs discussed “the most challenging part where we have not strengthened is the One Health approach”, a concept absent from FGD discourse. Conversely, FGD participants articulated barriers through lived experiences – the daily reality of inaccessible veterinarians, impassable roads during rainy seasons, and the economic desperation that drives self-medication and high-risk slaughter practices (S6 Appendix).

The absence of feedback mechanisms illustrated this divergence poignantly (S7 Appendix): KIIs discussed how unanswered cases “demotivate technical people” while FGD participants repeatedly stated, “we report, we never hear what happened”, expressing personal disillusionment rather than systematic analysis. Economic concerns similarly diverged – KIIs framed RVF as a threat to “international trade” and the national economy, whereas FGD participants focused on household-level impacts: “animals are used for paying dowry, so that means even people can’t marry”.

These complementary perspectives suggest that effective interventions must address both systemic barriers (identified by KIIs) and experiential barriers (identified by FGDs) simultaneously. Community-level sensitisation cannot succeed without addressing the transport and staffing gaps that leave reports unanswered; conversely, laboratory capacity building cannot compensate for the linguistic barrier of RVF having no local name, which prevents the disease from entering community discourse entirely.

### 3.3 Facilitators

**Table 3:** Facilitators for community-based abortion reporting, organised by the COM-B model.

| CAPABILITY | OPPORTUNITY | MOTIVATION |
| --- | --- | --- |
| **Psychological Capability**<br>Knowledge of standardised case definitions (IDSR)<br>Knowledge of RVF clinical signs among trained farmers<br>Understanding of reporting procedures.<br>Awareness of zoonotic transmission<br>**Physical Capability**<br>Presence of trained veterinary and health personnel<br>Technical skills of CHWs and district response teams<br>Diagnostic competence at national laboratories (UVRI, CPHL)<br>Presence of district task forces and rapid response teams<br>Analytical capacity of Emergency Operations Centre epidemiologists<br>Innovation capacity for diagnostic development | **Physical Opportunity**<br>Mobile phones & communication platforms (WhatsApp, radio)<br>Community health workers & local leaders (social resources)<br>National laboratory infrastructure (physical resources)<br>Emergency Operations Centre (EOC) (institutional resource)<br>Decentralised surveillance systems (structural resource)<br>Emerging diagnostic innovation platforms (technological resource)<br>Existing contact | **Reflective Motivation**<br>Experience from past outbreaks (collective memory)<br>Desire to access veterinary professionals.<br>Sense of civic duty to protect community health<br>Trust in government response systems<br>Perceived severity of RVF<br>**Automatic Motivation**<br>Fear of economic losses<br>Cultural obligations (dowry, bride price)<br>Fear of cultural disruption from livestock loss<br>Emotional response to |
|  | tracing systems (e.g., for TB)<br>• M-Trac platform (6767)<br>**Social Opportunity**<br>• Monthly LC1 meetings<br>• Community dialogues<br>• RVF priority disease status<br>• Community dialogues and sensitisation fora, NGO support (FAO, RedCross, Medical Teams International)<br>• Easy Communication between CHWs and Health Officers | visible disease severity |

#### a) Capability Facilitators

##### Psychological Capability Facilitators

i. **Knowledge of Standardised Case Definitions (IDSR):** The existence of standardised case definitions, particularly those described in the Integrated Disease Surveillance and Response (IDSR) guidelines, was reported as a foundational element for surveillance. A policymaker noted that RVF surveillance in humans used “the standardised case definition that is always applied” (KII-PM–8). This indicates that formal and consistent criteria for identifying and classifying disease events are established at a national level. These definitions provide a common language and framework for health personnel involved in surveillance activities.
ii. **Knowledge of RVF Clinical Signs in Animals:** Knowledge about the clinical signs of RVF in animals was frequently highlighted as a key facilitator, with 28 references in the codebook. Participants indicated that farmers who had received training were better able to identify abortions as a potential sign of RVF. This awareness of specific clinical signs, such as abortions and haemorrhagic symptoms, was reported to enable more accurate and timely reporting. This finding suggests that targeted education on disease recognition can significantly improve community-level surveillance capabilities.
iii. **Knowledge of RVF as a Zoonotic Disease:** Understanding that RVF transmits from animals to humans emerged as a key psychological capability (29 references). Participants demonstrated awareness that “this disease attacks both human beings and animals, but mostly people get it when they milk infected cows and drink unboiled milk or eat half-cooked meat” (FGD-LO-7, Rugaga). This knowledge creates a cognitive link between animal health and personal health, motivating reporting behaviour.

##### Physical Capability Facilitators

i. **Presence of Trained Veterinary and Health Personnel**: The presence of trained veterinary staff and health personnel, equipped with technical skills, was found to be a crucial asset. These individuals have the expertise required to understand and apply standardised case definitions, conduct investigations, and manage disease outbreaks. Their training enables them to perform specific tasks within the surveillance system, contributing to the overall capacity for disease detection and response.
ii. **Diagnostic Competence at National Laboratories (UVRI, CPHL):** National reference laboratories, such as the Uganda Virus Research Institute (UVRI) and the Central Public Health Laboratories (CPHL), were identified as possessing significant diagnostic capabilities. These facilities are equipped to perform advanced laboratory tests, including those necessary for confirming RVF. A policymaker mentioned, “an investigation form that is centrally located in the central public health laboratory and UVRI” (KII-PM-8), indicating that these laboratories serve as central hubs for sample analysis and confirmation. This centralised infrastructure provides the authoritative confirmation required for formal responses to disease reports.
iii. **Analytical Capacity of Emergency Operations Centre (EOC) Epidemiologists**: The Emergency Operations Centre (EOC) was described as a central coordination hub with epidemiologists possessing analytical capacity. This capacity involves the ability to process incoming information, verify reports, and coordinate responses. A member of the Ministry of Health technical staff stated: “…when we receive an alert in the emergency operations centre, then we verify the information” (KII-MOH-3). This indicates that the EOC has the personnel and processes in place to analyse surveillance data and trigger appropriate actions.
iv. **Presence of District Task Forces and Rapid Response Teams:** The existence of district-level coordination mechanisms was noted as a critical physical capability (5 references). A district official explained: Isingiro district has an existing district task force which, in times of outbreak like this, sits every week; it has an active district response team… the response capacity to survey is there” (KII-DTO-3). Another KII participant added: “we have fellows on the National Rapid Response Team that are actually capable of responding within 24 to 48 hours” (KII-MOH-13).
v. **Innovation Capacity for Diagnostic Development**: Uganda was found to have innovation platforms in the country dedicated to diagnostic design and development. A national technical officer highlighted this, stating: “We have an open platform where we can design a diagnostic using PCR. We also have a sequencing platform where we can sequence and be able to tell which genotypes are circulating” (KII-NTO-4). This indicates a sophisticated capability to develop and adapt diagnostic tools, including molecular methods such as PCR and sequencing, which allow the identification of specific pathogen genotypes. This capacity extends beyond routine diagnostics to include the ability to respond to emerging threats and evolving viral strains. This facilitates reporting behaviour.
vi. **Community Structures (CHWs, Local Leaders, District Rapid Response Teams**): The presence and active involvement of various community structures were identified as significant facilitators. These included Community Health Workers (CHWs), local council leaders (e.g., LC1 chairpersons), and District Rapid Response teams. These individuals and groups were observed to use existing communication platforms to bridge information gaps between local communities and higher authorities. A policymaker noted the role of CHWs as “foot soldiers put in the villages who could actually be given some information which they can easily pass on to the community to be able to look out for these signs” (KII-PM– 8). Similarly, a local council leader described the reporting pathway: “When there is a disease, we report to the chairperson of the LC 1 [Local Council], and they take it to the town council to the veterinary doctor” (FGD-LCL-17). These structures integrate surveillance within established community networks, providing accessible pathways for reporting.
vii. **Decentralised Surveillance System:** The surveillance system was described as having a decentralised structure, which was identified as a key facilitator. This structure includes focal points and task forces at the district-level. A National Technical Officer explained, “We have a surveillance team led by a focal person… it is the task force that supports them” (KII-NTO-5). This decentralisation places surveillance coordination closer to the community level, theoretically shortening the communication and response chain between communities and decision-makers.

#### b) Opportunity Facilitators

##### Physical Opportunity Facilitators

i. **Mobile Phones and Communication Platforms:** Relevant to both Capability and Opportunity. Communication platforms provide the physical infrastructure enabling reporting. A district official noted: “We use radio talk shows to pass on the message to those in far distances who need to know” (KII-DTO-18). This technological presence created physical pathways for information flow to new places or people.
ii. **Presence of Laboratory Hub System**: The existence of a functional laboratory hub system has been frequently cited (12 references) as a significant physical opportunity. This system includes a cold chain (7 references) for sample preservation and transport, and access to reference laboratories for advanced analysis (8 references). These infrastructures provide the necessary physical resources for diagnostic confirmation and support the overall surveillance efforts. Furthermore, the presence of research infrastructure (6 references) was noted as an opportunity to advance diagnostic capabilities.
iii. **Emergency Operations Centre as Institutional Resource:** Beyond its analytical capacity, the EOC functions as an institutional resource that coordinates multi-sectoral response. A KII participant noted: “I will credit the way we set up our surveillance systems, which is quite good now because when we receive an alert in the EOC, then we verify the information” (KII-MOH-18).
iv. **M-Trac Platform and Electronic Reporting:** The existing mobile-based reporting system for human health presents an opportunity for integration. A KII participant from FAO noted: “ We have been having the event mobile application reporting, that is M-Trac… Officers can now send in immediate mobile reports, whenever there is a problem; they can send a signal and reports, and the country can be alerted to where the problem is” (KII-NTO-5).

##### Social Opportunity Facilitators

i. **Existing Contact Tracing for other Endemic Diseases (e.g., Tuberculosis):** The established system for contact tracing, particularly for diseases such as tuberculosis (TB), was identified as a pre-existing opportunity that could be leveraged. A health technical staff member stated that “There are also some surveillance activities that are performed, and we could integrate the RVF, for example, we have what we call active case search” KII-DTO-3). This indicates that community members and health workers are already familiar with procedures for disease investigation and reporting, and platforms like M-Trac are in use for human health events.
ii. **Existing Indicator and Event-Based Surveillance Systems under the Ministry of Health:** The Ministry of Health operates a dual system of indicator-based and event-based surveillance, which was noted as a robust opportunity. A member of the Ministry of Health technical staff explained: “We have a robust system now in the country; we do both indicator-based surveillance and then event-based surveillance… alerts… come to the EOC, and then eventually we respond quite early” (KII-MOH-3). This signifies a mature surveillance infrastructure within the human health sector, capable of detecting and responding to health alerts.
iii. **Community Governance Mechanisms (LC1 Meetings, Community Dialogues):** Regular community gatherings, such as monthly meetings of the local council (LC1) and broader community dialogues, were identified as existing social opportunities. A livestock owner reported that, “…we meet every month and discuss and exchange knowledge. We meet there with the veterinarians and report” (FGD-LO-2). These forums provide routine and collective settings for information sharing and interaction between community members and local authorities, including veterinary personnel. Community sensitisation events (3 references) were also highlighted as a valuable opportunity to raise awareness.
iv. **RVF Being a Priority Zoonotic Disease:** The formal prioritisation of RVF as a zoonotic disease in Uganda was recognised as a significant policy opportunity. A member of the Ministry of Health technical staff stated: “…we supported joint risk assessment in 2019, and the country prioritised RVF among the three diseases…” KII-NTO-28). This prioritisation indicates that RVF is officially recognised as a legitimate concern at a high policy level, theoretically attracting political will, resource allocation, and inter-sectoral attention.
v. One Health Coordination and NGO Support: Improved One Health coordination (8 references) was identified as a social opportunity, particularly at the sub-national level, allowing for better integration of human and animal health efforts.

Support from non-government organisations (6 references) was noted as a valuable resource. A district official acknowledged: “Capacity building, trainings they were once supported by the Ministry of Health under Uganda Virus Research Institute, we had trainings from OXFAM, yes, trainings in the refugee settlements have been supported by Medical Teams International and Uganda Red Cross Society” (FGD-LCO-12). Easy communication between Village Health Teams (VHTs) and Health Officers (HOs) (3 references) was also highlighted as a facilitator of information flow.

#### c) Motivation Facilitators

##### Reflective Motivation Facilitators

i. **Experience from Past Outbreaks:** Memory and experience from previous RVF outbreaks were frequently cited (10 references) as a powerful motivator for reporting. A KII participant noted: “Preparedness Uganda I think has done so well in generally outbreaks since Ebola, which was recently contained in a short time… we have built a lot of experience in outbreaks, especially on the human side” (KII-MOH-19). A district official added: “When it comes to a human being, because you know, having had a background of COVID-19 and recently we had this Ebola alert, we strengthened the systems in the communities, so anything suspicious now the community responds very fast” (KII-DTO-25). This collective memory of disease events contributes to increased awareness and willingness to participate in surveillance activities.
ii. **Perceived Severity of RVF:** The perceived severity of RVF, particularly its haemorrhagic manifestations, was identified as a motivator. A MoH Technical Officer noted, “…The issue is the seriousness of the disease because it presents in unusual ways that people get scared, and once they get scared, they report…” (KII-MOH-5). This indicates that the visible and alarming symptoms of RVF, which can be frightening, prompt individuals to report cases.
iii. **Desire to Access Veterinary Professionals:** The accessibility of veterinary professionals was reported to be a key motivator for the reporting. A livestock owner stated that: “Also, the availability of vets is one that we can report to and get help from… they are easily accessible” (FGD-LO-1). This suggests that the perceived ease of reaching expert help and the physical proximity of veterinary services directly encourage reporting behaviour, as farmers anticipate receiving assistance and solutions for their animals.
iv. **Trust in Government Response Systems:** Trust in the government’s ability to respond effectively to disease outbreaks was identified as a motivational factor. When community members believe that reporting will lead to a timely and appropriate response from authorities, their willingness to engage with the surveillance system increases.
v. **Sense of Civic Duty to Protect Community Health:** It was observed that the sense of civic duty to protect community health contributes to the motivation to report. This suggests that some individuals are driven by a broader responsibility to safeguard the well-being of their community, extending beyond personal economic concerns.

##### Automatic Motivation Facilitators

i. **Fear of Economic and Cultural Loss:** Fear of economic and cultural losses was found to be a powerful motivational driving force for reporting. Livestock owners expressed concern about “significant economic losses due to death and abortion” (FGD-LO-13). This economic anxiety was compounded by cultural considerations, as an official explained: “Animals are used… to pay the dowry… which means that… people cannot marry… because there is a quarantine” (KII-MOH-7). This indicates that the potential loss of livestock impacts not only financial well-being but also deeply embedded social and cultural practices.
ii. **Fear of Cultural Disruption:** Closely related to economic loss, the fear of cultural disruption emerged as a distinct automatic motivator. Participants recognised that livestock loss affects social standing, marriage arrangements, and community relationships – creating powerful emotional responses that bypass reflective deliberation and prompt immediate action

#### 3.3.2 Barriers

**Table 4:** Barriers to community-based abortion reporting, organised by the COM-B model.

| CAPABILITY | OPPORTUNITY | MOTIVATION |
| --- | --- | --- |
| **Psychological Capability**<br>Poor knowledge of RVF clinical signs among farmers (55 references)<br>Confusion of RVF with other diseases (14 references)<br>Lack of skills to distinguish normal from abnormal abortions (4 references)<br>Insufficient training on reporting procedures (1 reference)<br>**Physical Capability**<br>Few veterinary staff, big district (understaffing – 18 references)<br>Limited diagnostic expertise at | **Physical Opportunity**<br>Inadequate transport infrastructure (13 references)<br>Limited district laboratory capacity (physical resource gap) (7 references)<br>Rough terrain and geographic remoteness (9 references)<br>Poor coordination between One Health sectors (structural barrier)<br>Absence of formal | **Reflective Motivation**<br>Absence of feedback mechanisms after reporting (12 references)<br>Fear of Quarantine and movement restrictions<br>Distrust in government response<br>Low perceived risk of RVF during inter-epidemic periods<br>Economic sensitivity of RVF reporting (fear of trade/tourism impacts) |
| district level<br>• Weak policies and regulations (8 references)<br>• Underpayment of animal health workers (1 reference)<br>• Low field/farm visits by para-veterinary staff<br>• Limited cold chain/storage capacity (4 references)<br>• Underutilisation of people at grassroots/community level | feedback mechanisms<br>**Social Opportunity**<br>• Poor One Health coordination at sub-national level (7 references)<br>• Politics and public opinions (7 references)<br>• Traders' fear of market disruption (5 references)<br>• RVF not prioritised in practice<br>• No known RVF name (1 reference)<br>• Two ministries in charge (MAAIF, MOH) creating coordination gaps.<br>• Corruption of leaders<br>• Limited research about RVF<br>• Poor communication about the disease (RVF) | • Lack of perceived value from reporting due to absence of feedback (demotivation)<br>• Preference for self-treatment practices over formal reporting<br>**Automatic Motivation**<br>• Fear of economic repercussions overriding civic duty (5 references)<br>• Emotional response to potential trade restrictions (1 reference)<br>• Despair from repeated responded reports (1 reference)<br>• Cultural shame associated with disease outbreaks (1 reference) |

##### a) Capability Barriers

###### Psychological Capability Barriers

i. **Poor Knowledge About RVF:** The most frequently cited barrier to community reporting, with 55 references, was poor knowledge about RVF among community members. This fundamental lack of understanding significantly impedes the ability to recognise and report potential cases. The participants noted that farmers often studied RVF the last time in school and were only reminded during outbreaks. A key informant from academia stated: “What stops us to curb down the speed of this disease is limited knowledge about the disease” (FGD-LO-5). Another academic key informant further elaborated on the community-level capacity: “The problem I see from the district downwards, which we call the community-level; capacity is low. So, in other words, we have capacity upwards, but we don’t have capacity downwards because the farmers do not know the disease, they can’t report it.” (KII-ACAD-1). This lack of recognition was so profound that “All the RVF cases that we have confirmed have been submitted as suspects of Ebola, not Rift Valley fever.” (KII-ACAD-2). This indicates that communities cannot report what they do not recognise, leading to misidentification of diseases.
ii. **Confusion of RVF with Other Diseases:** A related and frequently mentioned barrier (14 references) was the confusion of RVF with other animal and human diseases. Farmers often struggled to differentiate RVF symptoms from those of other common animal diseases. Similarly, health workers sometimes confuse RVF symptoms in humans with malaria. This confusion was exacerbated by the absence of a local name for RVF, as highlighted by a key FAO informant. “Most people don’t know; the disease doesn’t have a local name. When you say RVF, it sounds new, and it can be confused with fever or malaria. So, there is still confusion” (KII-FAO-1). This linguistic and diagnostic ambiguity contributes to underreporting and misreporting.
iii. **Lack of Skills to Distinguish Normal from Abnormal Abortions:** Participants expressed difficulty in differentiating between normal, sporadic abortions and those that might indicate a disease outbreak. FGD participants repeatedly stated, “We don’t know if every abortion needs to be reported” (FGD-LCL-3). This suggests a gap in practical skills for discerning reportable events from routine occurrences.
iv. **Insufficient Training on Reporting Procedures:** Community members and some local-level personnel reported insufficient training on the specific procedures to report livestock abortions. This includes a lack of clarity on who to report to, what information to provide, and the expected follow-up process. Poor reporting mechanisms (5 references) were also noted as a barrier.

###### Physical Capability Barriers

i. **Few Staff, Big District:** The scarcity of veterinary and surveillance personnel, particularly in relation to the large geographical areas they cover, was a significant barrier to physical capability (18 references). Participants frequently highlighted that veterinary doctors often cover four or more sub-counties, leading to long response times. A participant in a Kitagati FGD explained: “A veterinary officer looks after like 4 sub-counties. For example, like now here, I can spend like 5 months without seeing the doctor. Now that person, when will you report to him that cows are sick?” (FGD-LO-2). The issue was compounded by the lack of replacements for retired personnel. “We have this disease, but we do not have a doctor in our sub-county of Kashumba. We need a doctor to come and educate us. That is a major challenge that we have. The doctor who was there retired.” (FGD-LO-5). This shortage of personnel directly affects the ability to respond quickly and investigate.
ii. **Limited Diagnostic Expertise at District Level:** Diagnostic expertise was reported to be limited at the district level. This means that local veterinarians or health personnel may not have the specialised knowledge or equipment required to accurately diagnose RVF or other causes of abortion without relying on central laboratories.
iii. **Underpayment and Poor Facilitation of Staff:** Staff motivation was undermined by inadequate facilitation (6 references) and low pay (1 reference). A KII participant stated: “Some laboratory personnel, sometimes they can get a suspect or probable case and fail to work on samples or screening… because these people are not facilitated; they say why should I go ahead to get these samples when I am not facilitated” (KII-DTO-18). A FGD participant observed: “The challenges we face in fighting the RVF are that the veterinary doctors are not well paid, which means they are not motivated to work (FGD-LO-4, Rwetango). This creates a cycle where demotivated staff provide poor services, further discouraging community reporting.
iv. **Weak Policies and Regulations:** Weaknesses in policies and regulations (8 references) were identified as a capability barrier, indicating a lack of clear frameworks or enforcement mechanisms to support effective surveillance.
v. **Farmers’ Self-Diagnosis and Management:** The common practice of farmers who engage in self-diagnosis and the management of animal diseases (7 references) was noted as a barrier, as it bypasses formal reporting channels and professional veterinary care.

#### b) Opportunity Barriers

##### Physical Opportunity Barriers

i. **Inadequate Transport Infrastructure:** The lack of adequate transport infrastructure was identified as a critical barrier. A district technical officer stated, “Our surveillance personnel lack transport means… This specific vehicle may be needed for other surveillance activities” (KII-DTO-10). This indicates that the personnel responsible for follow-up investigations and sample collection often lack reliable means of transportation, which hinders timely responses. District officials also noted that “we share one vehicle between human and animal health, and it is often broken” (KII-DTO-12). This issue of limited transportation means was frequently mentioned (13 references), with vehicles often being called back mid-surveillance, making the process inefficient.
ii. **Limited District Laboratory Capacity (Physical Resource Gap):** Diagnostic capacity was reported to be severely limited at the district level. A MoH Technical Officer noted: “…not having access to a state-of-the-art lab that can easily conduct abortion screening is also a barrier…” (KII-MOH-8). This indicates a lack of physical laboratory infrastructure and equipment at the local level, making it difficult to conduct timely diagnostic tests for livestock abortions. This often results in the reliance on distant national laboratories, leading to delays. Other physical resource gaps included limited storage capacity (4 references) and general limited infrastructure (4 references).
iii. **Geographic Remoteness and Challenging Terrain:** The geographic remoteness of certain communities was reported as a barrier. This physical distance from veterinary services, health facilities and transport routes makes it difficult for community members to report cases and for response teams to reach affected areas promptly. Furthermore, rough terrain (9 references) was frequently cited as an additional physical obstacle, making access to remote areas challenging for surveillance teams. The sheer size of the districts (5 references) also contributed to the difficulty in covering all areas effectively.
iv. **Large District Size for Surveillance Coverage:** The sheer size of the districts (5 references) contributed to the difficulty in covering all areas effectively. A district official noted: “Isingiro is a very big district, very many roads are full of mud and distant, by the time you reach there, you find that even this person you are going to take samples from, he has already been told by people that we are coming, and sometimes they run away (KII-DTO-29). This geographical challenge compounds staffing shortages.
v. **Limited Cold Chain and Storage Capacity:** Limited storage capacity (4 references) and general limited infrastructure (4 references) were not cited as physical resource gaps. A FGD participant from Kikagati noted: “We were looking at other facilities for keeping drugs. We would fail to get them to keep up with vaccines. We did not have them. Even storage channels. Even when there is an outbreak, they use health centre III refrigerators to keep the vaccines” (FGD-LCO-Kikagati). A district official confirmed: “We have a challenge of storage facilities for keeping our drugs and vaccines as a department of veterinary” (KII-DTO-9)

##### Social Opportunity Barriers

i. **Poor Coordination Between One Health Sectors at Sub-National Level:** Poor coordination between human and animal health sectors, particularly at the sub-national (district) level, was a pervasive barrier. A MoH Technical Officer observed: “…the most challenging part where we have not strengthened is the ‘one health approach’…” (KII-MOH-5). This indicates a lack of operational collaboration and resource sharing between the two sectors, leading to fragmented efforts in surveillance and response.
ii. **Political and Public Opinion Factors**: Political considerations and public opinion (7 references) were identified as social opportunity barriers. These factors can influence decision-making and resource allocation, potentially hindering surveillance efforts. The fear among traders of market disruption (5 references) due to RVF reports also created a disincentive for transparent reporting. Additionally, the perception that RVF was not sufficiently prioritised (3 references) and the involvement of two ministries in charge (2 references), leading to potential bureaucratic hurdles, were noted as structural barriers.
iii. **No Local Name for RVF (Linguistic/Cultural Barrier):** The absence of a local name for RVF was identified as a significant barrier to recognition and reporting. A policymaker stated that “Most people do not know the disease [RVF], as it does not have a local name…” (KII-PM-1). This linguistic gap prevents the disease from entering common community discourse and shared knowledge, making it difficult for community members to communicate about or identify the disease.
iv. **Economic Sensitivity of RVF Reporting (Fear of Trade/Tourism Impacts):** The economic sensitivity surrounding RVF reporting was identified as a major barrier. A district official candidly stated, “When you talk about the Rift Valley fever, you spoil someone’s market for products… as a country” (KII-DO-3). This indicates a fear among officials and potential community members that reporting RVF could lead to negative economic consequences, such as trade restrictions or impacts on tourism, creating disincentives for transparent reporting.
v. **RVF not prioritised in practice:** Despite policy-level prioritisation, the perception that RVF was not sufficiently prioritised in practice (3 references) was noted as a structural barrier. A KII participant from the Science, Technology and Innovation Secretariat – Office of the President (STI-OP) noted: “The opportunities for RVF, of course, as other diseases exist; the only unfortunate bit is that the country has not prioritised it” (KII-NTO-12). This gap between policy rhetoric and resource allocation undermines surveillance efforts.
vi. **Two Ministries in Charge:** The involvement of two ministries (MAAIF and MoH) (2 references) was noted as a potential source of bureaucratic hurdles and coordination gaps. A KII participant explained: “The challenge we have as a country with RVF is that it falls into two Ministries; we have MAAIF, MoH, because the disease starts in animals and small ruminants; it is detected late in most cases, a spill over (KII-ACAD-2). This structural fragmentation created ambiguity about leadership and accountability.
vii. **Limited Research about RVF:** Limited research (1 reference) was noted as a barrier to understanding disease drivers and designing evidence-based interventions. A KII participant noted: “There is also limited research because we don’t know the drivers of these outbreaks, the exact drivers explaining why the outbreaks have come up somewhere are lacking” (KII-NTO-2).

#### Motivation Barriers

##### Reflective Motivation Barriers

i. **Absence of Formal Feedback Mechanisms:** The lack of formal mechanisms for providing feedback to reporters was consistently cited as a barrier. This means that people who report cattle abortions often do not receive information on the outcome of their report, such as diagnostic results or actions taken. The lack of feedback after making a report was the most cited demotivating factor. A policymaker observed that “It has not been easy because when someone reports, they should get action and feedback… most technical people have been demotivated” (KII-PM-2). Participants in various FGD also stated that “we report, but we never hear what happened” (FGD-LO-3, FGD-LCL-7, FGD-LO-11). This indicates that when reporting efforts do not produce visible results or communication, individuals perceive their actions as futile, leading to reduced motivation to report in the future.
ii. **Distrust in Government Response:** Distrust in the government’s ability or willingness to respond effectively and fairly was identified as a motivational barrier. When community members lack confidence in authorities, they are less likely to participate in formal reporting systems.
iii. **Preference for Self-Treatment Practices Over Formal Reporting:** The common practice of self-treatment for animal illnesses has been reported to be a significant barrier to formal reporting. A livestock owner explained, “What stops people from reporting… People were used to buying drugs in stores, so they decided to buy from stores and treat their animals themselves” (FGD-LO-6). This indicates that community members often opt for readily available, informal solutions, bypassing the formal surveillance system.
iv. **Fear of Quarantine and Movement Restrictions:** A significant motivational barrier was the fear that quarantine and movement restrictions would be imposed after an RVF report. This fear stems from the potential economic disruption and social inconvenience associated with such measures, leading people to hesitate to report.
v. **Low Perceived Risk of RVF During Inter-Epidemic Periods:** During periods between RVF outbreaks (inter-epidemic periods), the perceived risk of the disease was reported to be low. This reduced perception of threat leads to decreased vigilance and motivation to report potential cases, as the urgency and immediate threat are not felt.

##### Automatic Motivation Barriers

i. **Economic Sensitivity of RVF Reporting (Fear of Trade/Tourism Impacts):** As noted in Opportunity Barriers, the economic sensitivity of RVF reporting also functions as a strong motivational barrier. Fear of negative economic repercussions, such as trade bans or reduced tourism, directly discourages individuals and even officials from reporting suspected cases, as they anticipate adverse outcomes for themselves or the country.
ii. **Despair from repeated Unanswered Reports**: Beyond the reflective assessment that reporting is futile, repeated experiences of unanswered reports create an automatic emotional response of hopelessness and resignation. Participants who had reported multiple times without any feedback or action described a sense of learned helplessness – they simply stopped reporting because experience had taught them it made no difference.
iii. **Cultural Shame associated with Disease Outbreaks:** Implicit in several responses was a sense of cultural shame or stigma associated with having a disease in one’s herd. This emotional response can automatically suppress reporting, as farmers may fear being seen as having “unclean” animals or being responsible for bringing disease into the community.

## Discussion

This study provides a theoretically grounded analysis of the behavioural determinants shaping community-based livestock abortion reporting for RVF surveillance in Uganda. By applying the COM-B framework and systematically comparing perspectives across participant groups, we have generated three novel insights that extend current understanding of zoonotic disease surveillance in resource-limited settings. First, the barriers and facilitators are not merely multiple, but they are interconnected; deficits in one domain amplify deficits in others.

Secondly, the same phenomenon is understood fundamentally differently by policymakers and communities, with implications for intervention design that addresses both systemic structures and lived experiences. Thirdly, the ranking of barriers by frequency and emphasis reveals that the most powerful demotivator was the absence of feedback, which was different from the most cited barrier – knowledge deficits. This suggests that intervention priorities must be guided by impact rather than prevalence alone.

The most frequently cited barrier, poor community knowledge about RVF (55 references), represents a foundational deficit that cascades through the entire surveillance system. Communities cannot report what they cannot recognise, especially a disease that lacks a local name. The finding that all confirmed RVF cases were initially submitted as Ebola suspects illustrates the profound consequences of this cognitive gap. This aligns with Tumusiime et al (Tumusiime et al., 2023b), who documented similar knowledge deficits in Ugandan pastoral communities, and we extend their work by quantifying the magnitude of the gap and revealing its linguistic dimension. Although Tumusiime et al found that pastoralists in Napak district had a local name for RVF (*Lonyang* – a term symbolising jaundice, high fever, abortions in pregnant cows and sudden deaths in calves, this name was not reported in Isingiro district. Their study sampled from four districts of Napak, Butebo, Isingiro and Lyantonde and reported that livestock keepers in all four districts had knowledge of RVF and even had local names or descriptions of it; the actual published findings do not specify the local name for Masha (sub-county), one of their study sites. The persistence of knowledge gaps is corroborated by multiple studies across East Africa. Chiuya et al (2023) demonstrated that in Baringo South, Kenya, only 9.6% attained at least half of the total knowledge score on clinical signs, transmissions, and prevention (Chiuya et al., 2023). Odinoh et al (2025) in their multi-country study in Uganda, the Democratic Republic of Congo and Kenya, found that only 20.5% of 4,806 participants had basic knowledge of RVF (Odinoh et al., 2025). The poor community knowledge of RVF and the absence of a local name for RVF is not merely a translation problem but a structural constraint. A participatory process that enables communities to name and conceptualise the RVF threat in their own terms would contribute to addressing this gap.

The second-ranked barrier, severe understaffing of veterinary professionals (18 references), represents a systemic gap. The finding that veterinary officers and paraveterinary staff cover up to four or more sub-counties and that the district administrators are unaware of true staffing levels reveals gaps in human resource governance that no amount of community sensitisation can overcome. This aligns with Vudriko et al (Vudriko et al., 2021) and Kebede et al (Kebede et al., 2025), who documented identical challenges across East Africa, but our findings add a crucial dimension: the staffing gap is uniformly distributed but concentrates its impact in remote communities, where farmers report waiting “5 months without a veterinary doctor”. Indeed, this was reported in our broader study, where we engaged closely with the private veterinary practitioners in Isingiro district, which facilitated reporting and investigation of livestock abortions (Walekhwa et al., 2025). This geographical inequity means that surveillance operates as a two-tier system: Response in accessible areas is present, but is absent in hard-to-reach locations, where ecological conditions may heighten disease risk. Addressing this requires not simply increasing the absolute numbers of staff, but targeted deployment to underserved areas with appropriate retention incentives (Driscoll, 2022).

The third-ranked barrier was the absence of feedback (12 references), which was perhaps the most striking finding, revealing that motivational factors can outweigh capability deficits in determining reporting behaviour. Despite being cited less frequently than knowledge gaps, the absence of feedback emerged as the most powerful demotivator across all participant groups. The repeated refrain “we report, but we never hear what happened” represents not merely a request for information but a fundamental breakdown in the social contract between communities and the surveillance system. When reporting yields no visible response, farmers rationally conclude that their effort is futile and revert to self-treatment practices. This finding resonates with Carlfjord et al.(Carlfjord et al., 2018) and Chilundo et al (Chilundo et al., 2004), who documented similar disengagement in health incident reporting systems when feedback was absent. In contrast, the issue of reporting needs to be viewed as a potential barrier. How? If farmers report and the feedback from the government is to institute geographical quarantine or livestock animal movement restriction, this is likely to hinder future reports (Ilukor et al., 2022; Mubiru et al., 2023b). This is because farmers rely on livestock for their livelihoods, and quarantining livestock movements means limited or no trade, likely shutting their businesses. One possible intervention for this is coming up with clear and stronger compensation mechanisms for the farmers. However, this compensation addresses the economic needs of the farmers, but livestock to farmers is beyond money. There are strong social ties to this, and it needs to be considered as well in the compensation plans/mechanisms. Future work needs to dismantle this if reporting for surveillance is to be scaled up effectively. Crucially, our comparative analysis reveals that the absence of feedback operates differently across levels: policymakers experience it as professional demotivation, whereas communities experience it as personal disillusionment. Effective feedback mechanisms must therefore operate at both levels, providing technical case updates to officers while offering communities acknowledgement and explanation (Mogg, 2022).

The systematic comparison between KII and FGD perspectives reveals that barriers are not simply present or absent but are understood and experienced differently across levels of the system. This divergence is not a limitation to be resolved but an analytical asset that illuminates the multi-level nature of surveillance failure. Policymakers frame economic concerns at the macroeconomic level (threats to international trade, national tourism) while communities experience them at the household level (inability to pay school fees, loss of dowry cattle). Policymakers discuss One Health coordination failures as institutional problems; communities experience their effects as inaccessible veterinarians and unresponsive systems. This implies that top-down solutions developed from policy perspectives alone will miss the experiential realities that determine whether communities use surveillance systems or not. Bottom-up solutions addressing immediate community frustrations will fail if they do not tackle the structural causes that perpetuate those frustrations. Effective intervention requires integrating both perspectives, addressing understaffing while simultaneously ensuring remaining staff are accessible, establishing feedback systems while ensuring they provide meaningful acknowledgement (Michie et al., 2011; Mogg, 2022).

Our findings provide empirical validation of the theoretical COM-B interconnectedness that Michie et al. posited, but few studies have demonstrated it in practice. The absence of feedback (opportunity failure) was the most powerful demotivator (motivation failure), creating a vicious cycle where disengaged reporters never develop the capability that comes from sustained engagement. Conversely, fear of economic loss (automatic motivation) drove reporting even where knowledge was poor (capability deficit), demonstrating that motivation can partially compensate for capability limitations. However, this compensatory mechanism is fragile – when motivated reporters encounter an unresponsive system, their motivation rapidly erodes, and they revert to self-treatment. This explains why training interventions alone consistently fail to produce sustained behaviour change (Ssengooba et al., 2012). Knowledge without a responsive system to receive reports and provide feedback is like a seed without soil. The seed may germinate briefly but cannot take root. Interventions must therefore address all domains simultaneously, recognising that gains in one domain will be lost if deficits in others are not addressed.

With these findings, we suggest four priority areas for interventions: First, address the foundational knowledge deficit through participatory processes that co-develop local RVF terminology, integrate disease recognition into existing community structures (LC1 meetings, VHT/CHWs networks), and leverage mobile phone penetration for ongoing reinforcement. Second, resolve the human staffing gaps through targeted recruitment for underserved areas, retention incentives, and engagement of private veterinary practitioners who are already serving communities but lack formal surveillance links. Third, establish simple, reliable feedback mechanisms that operate at both professional and community levels. For example, SMS notifications confirming report receipt, community meetings updating on investigation outcomes, and regular communications from district veterinary offices. Fourth, mitigate economic disincentives through compensation schemes, subsidised veterinary services for reported cases, or micro-insurance that transforms reporting from a financial threat to tangible benefits.

Crucially, these interventions must be designed and implemented through structures that already function, such as LC1 meetings, VHT networks, and WhatsApp groups, rather than creating parallel systems that communities must learn anew. The facilitators our study identified were: trained community health workers, mobile phone penetration, community governance structures, and national laboratory capacity, which provide ready-made infrastructure. This challenge is not a lack of capacity but a fragmentation of capacity across sectors and levels. Strengthening surveillance requires not building new systems but connecting and reinforcing existing ones.

This study possesses several strengths. The application of COM-B provides a systematic, theory-driven analysis that identifies modifiable behavioural determinants. The large, diverse sample and achievement of data saturation ensured comprehensive capture of relevant perspectives. The rigorous analytical approach, including independent coding verification and member checking, enhanced trustworthiness. The comparative analysis between participant groups adds methodological depth, revealing how the same phenomenon is understood differently across levels of the system. However, some limitations warrant consideration. As a qualitative study in a single district, findings may not be generalisable, though theoretical transferability is enhanced by the COM-B framework and consistency with regional literature. Social desirability bias may have influenced responses, particularly in FGD settings. The ranking of barriers and facilitators, while grounded in frequency and thematic emphasis, involves subjective judgment. The study captured processes at a single time point following a training intervention; longitudinal research would be needed to assess how barriers evolve. Finally, the study focused on barriers and facilitators to reporting and understanding the full surveillance cascade, which requires additional research.

## Acknowledgements

We are grateful to the technical staff at the Ministry of Agriculture, Animal Industry and Fisheries (Drs. Robert Mwebe and Dan Tumusiime) and the Isingiro District Local Government. We thank Conservation and Ecosystem Health Alliance (CEHA) for administrative and technical support during the implementation of the research. Special appreciation to Mellon Ainembabazi, who was very helpful in conducting the FGDs in the local language and later transcribing the transcripts. Lastly, we are grateful to the different leaders and farmers who agreed to participate in our FGDs and KIIs.

## Author contributions

AWW, AJKC, JLNW and LM conceptualised the study, developed study protocols and secured ethical approvals. AJKC mobilised funds to implement this study. AWW coordinated field activities and collected data. LNN and BN supported the data analysis. AWW drafted the first version of the manuscript. SAB, BN, and LNN reviewed and gave technical contributions to the manuscript. All authors reviewed and approved the final version of the manuscript.

## Data Availability Statement

All data are in the manuscript and/or supporting information files, which have all been included in this submission, and these have unrestricted access. For more information regarding this study, feel free to contact the corresponding authors at

## Competing Interests

All authors declare no competing interests.

## Funding Information

We are grateful for funding for this fieldwork from the Biotechnology and Biological Sciences Research Council (BBSRC) Impact Acceleration Account award, the Public Engagement Starter Grant, and the Nigeria Travel Grant from the University of Cambridge Centre for African Studies. The Alborada Trust supported AJKC and JLNW. The funder provided support in the form of salaries for authors AJKC and JLNW, but did not have any additional role in the study design, data collection and analysis, decision to publish, or preparation of the manuscript. The specific roles of these authors are articulated in the ‘author contributions’ section. AWW was supported by the Cambridge Trust for his doctoral studies at the University of Cambridge, United Kingdom.

### S1 Appendix: The Training Material used for sensitising different stakeholders about 1300 Rift Valley fever disease and the need for abortion surveillance

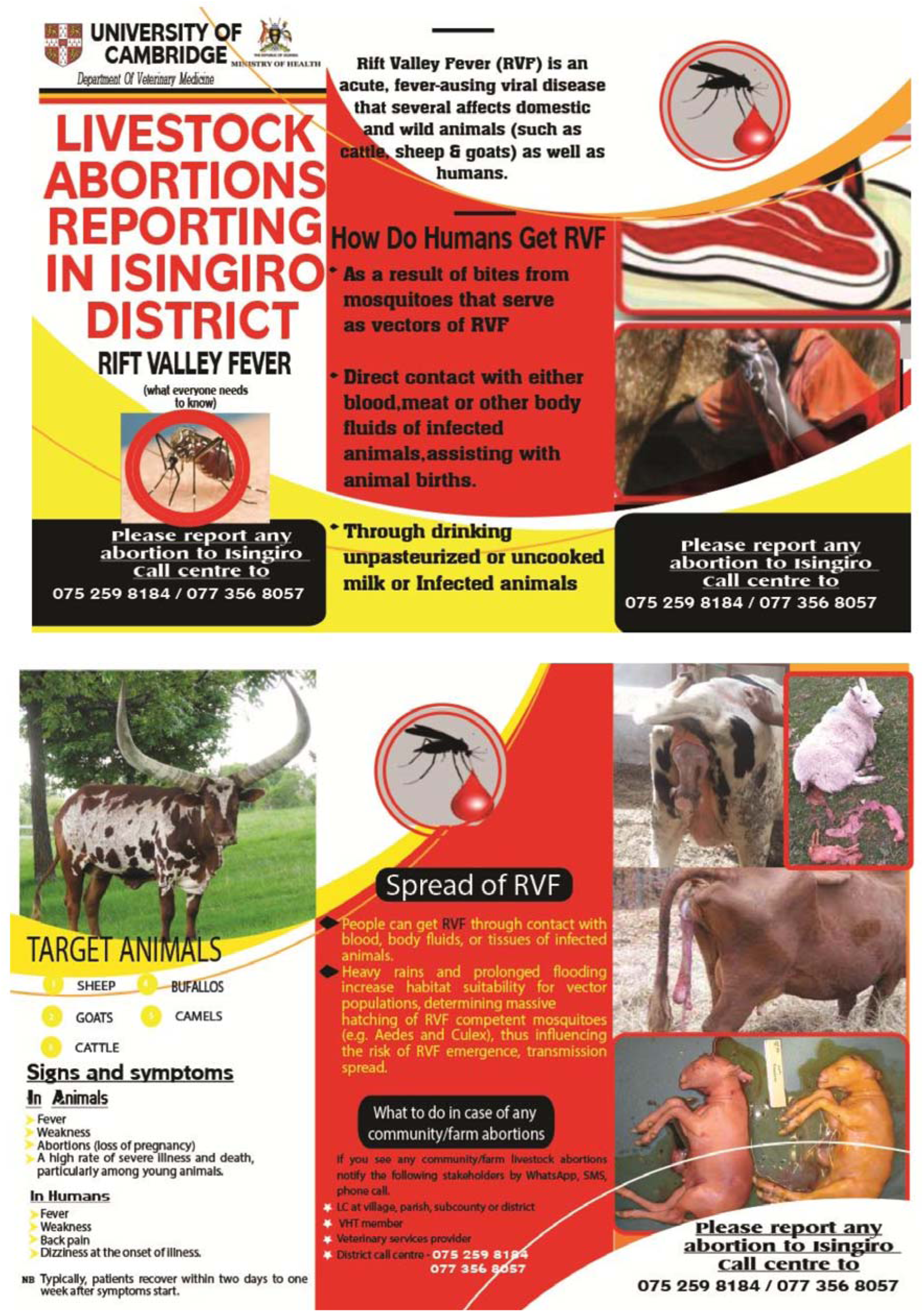

**S1 Table:** A List of participants trained as part of capacity building efforts.

| Category | Details of participants | Number |
| --- | --- | --- |
| District officials | District Health Officer, District Veterinary Officer, Assistant District Health Officer – Environmental Health, Assistant District Officer – Maternal Child Health, District Laboratory Focal Person, Senior Health Educator, Senior Environmental Health Officer | 7 |
| Sub county Technical Officers | Subcounty chief/Town Clerk, Health Inspector/Health Assistant, Veterinary Officer/Animal Husbandry officers, Community Development Officers, Parish Chiefs/Town Agents | 70 |
| Community Health Workers | Village Health Team members, Community Animal Health Workers | 35 |
| Livestock-engaged community members | Livestock owners, abattoir staff, local council leaders, diary staff | 788 |
| **Total** |  | **900** |

### S2 Appendix: The FGD guide used during Focus Group Discussions

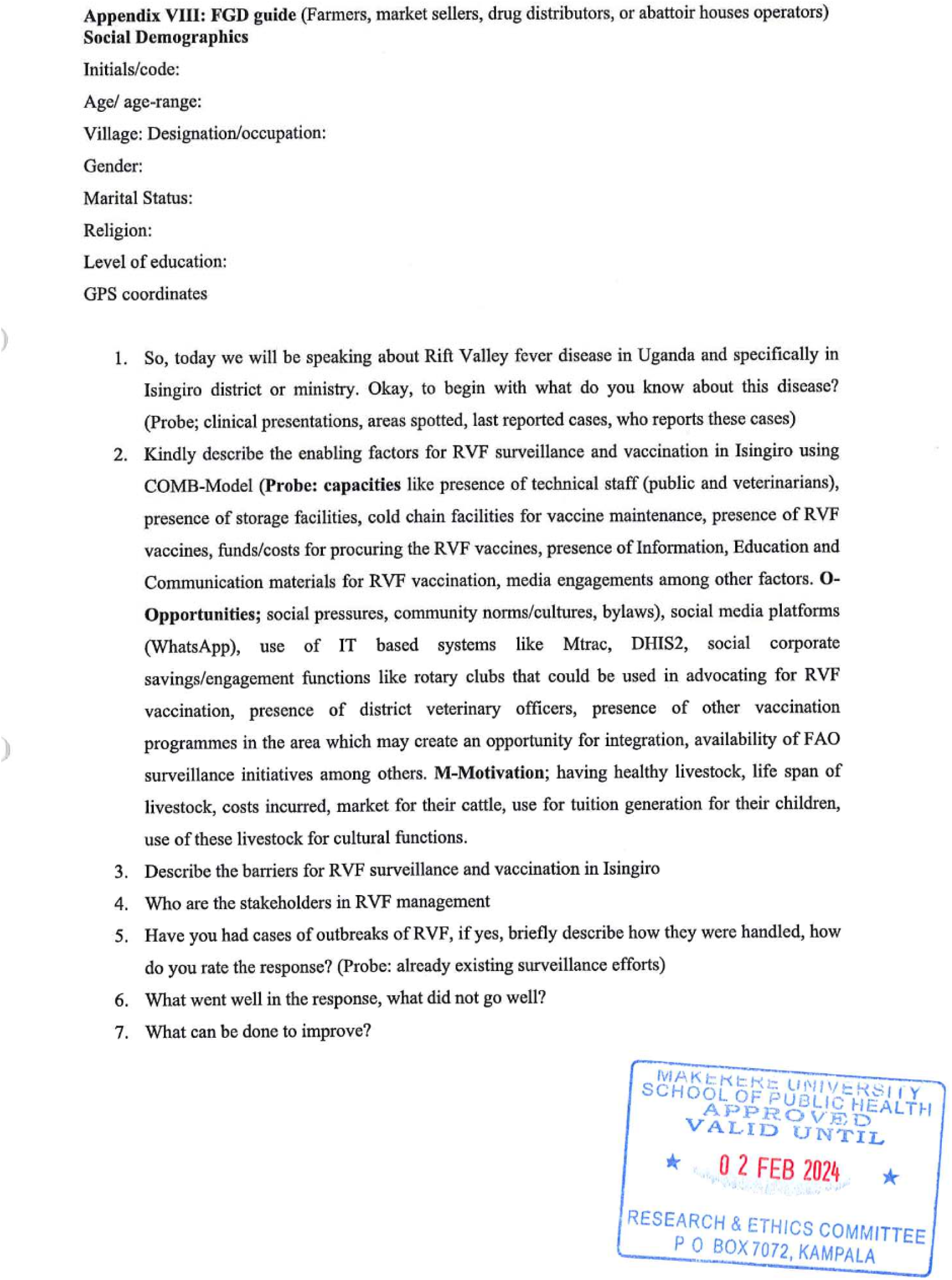

### S3 Appendix: The Key Informant Guide used for Key informant interviews

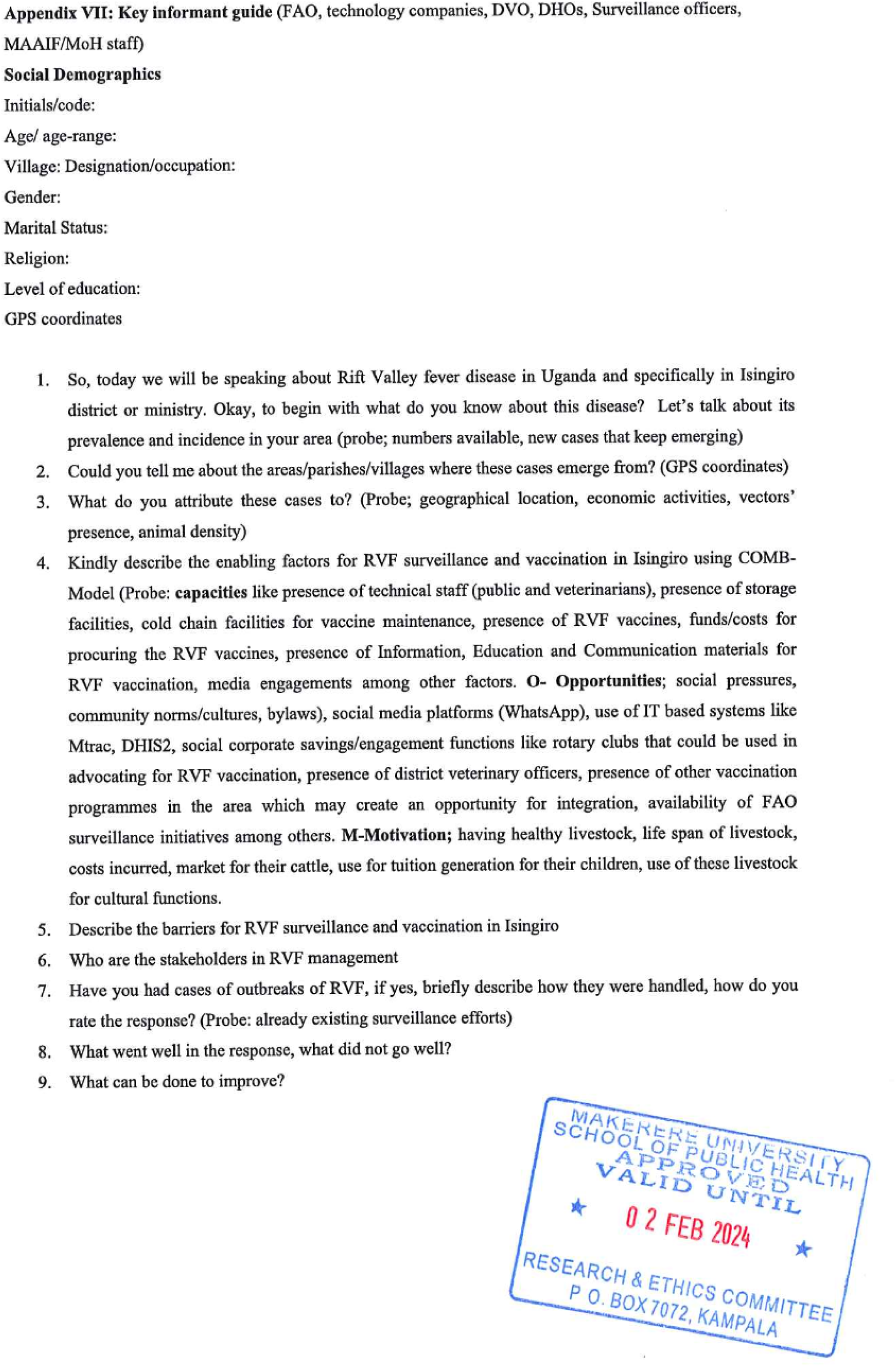

### S5 Appendix: Comparative Analysis and Ranking of Facilitators and Barriers: KII vs FGD Perspectives

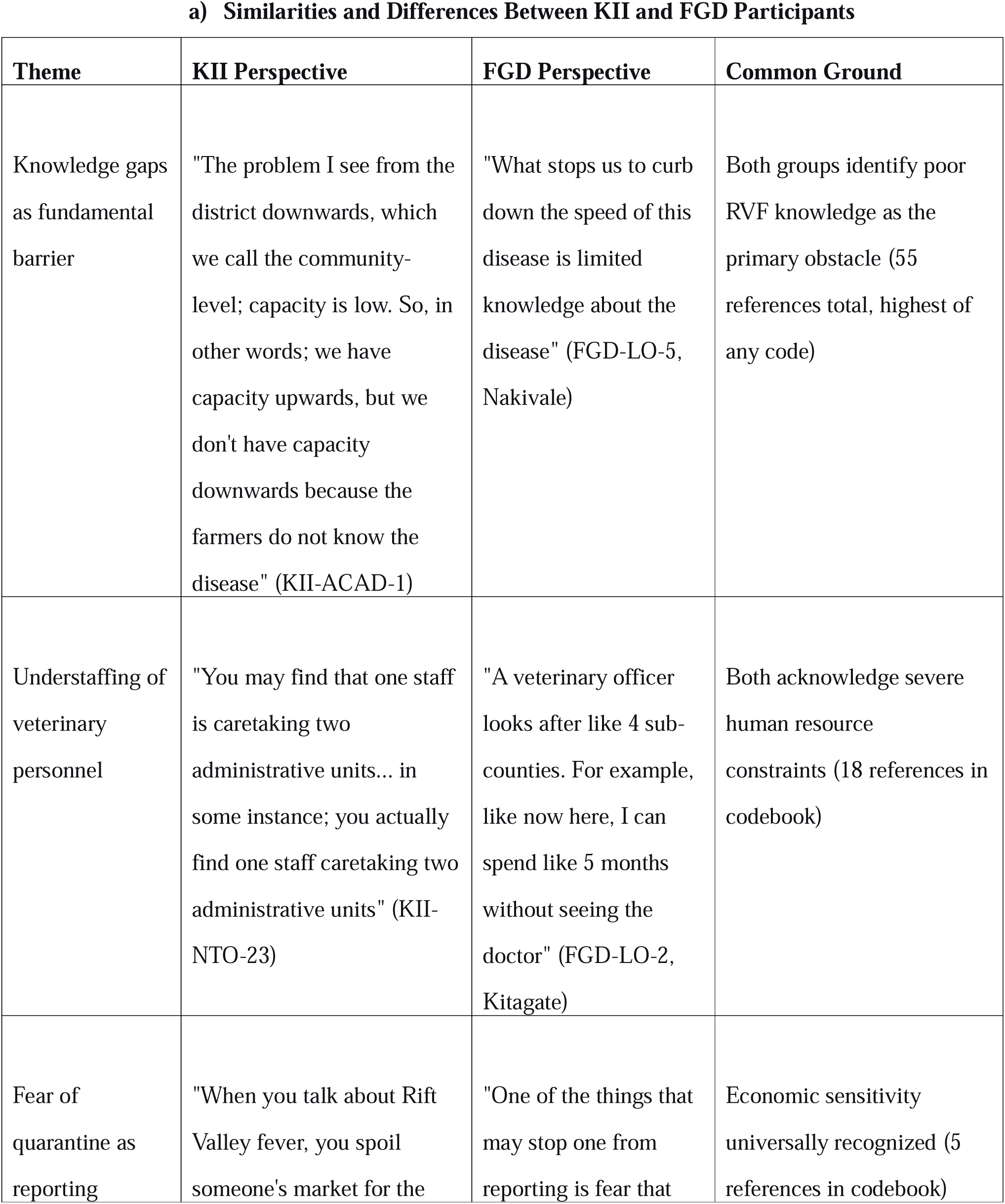
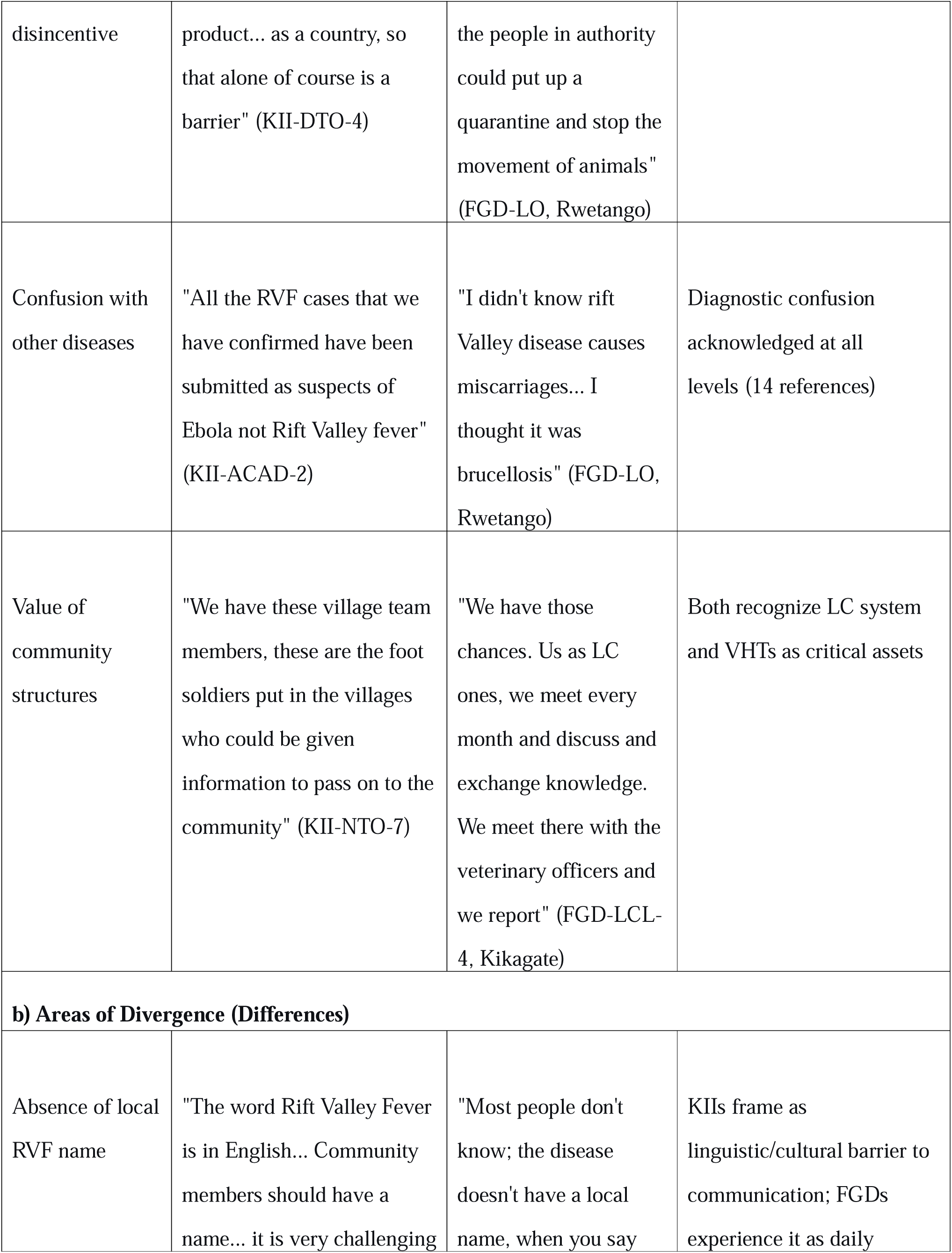
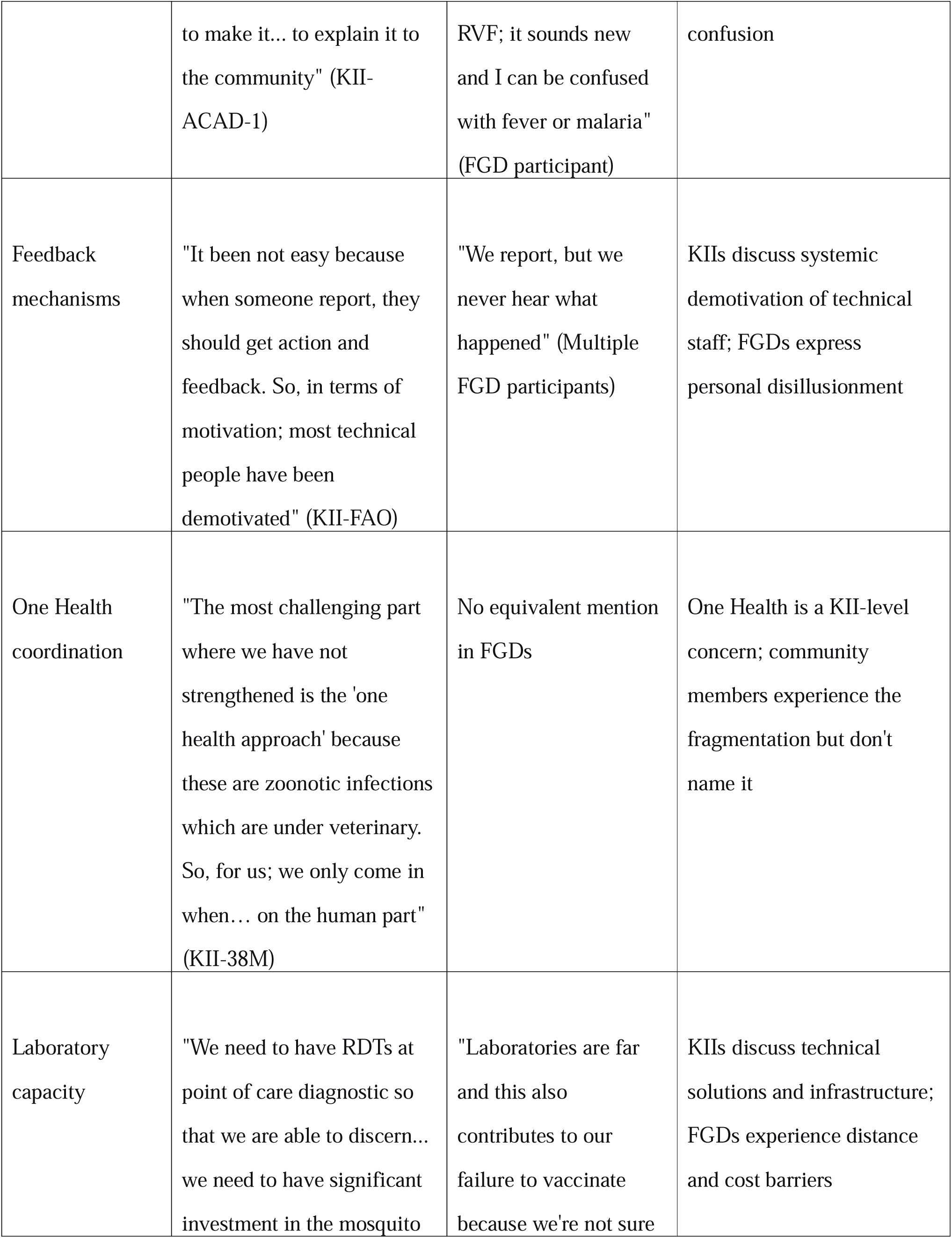
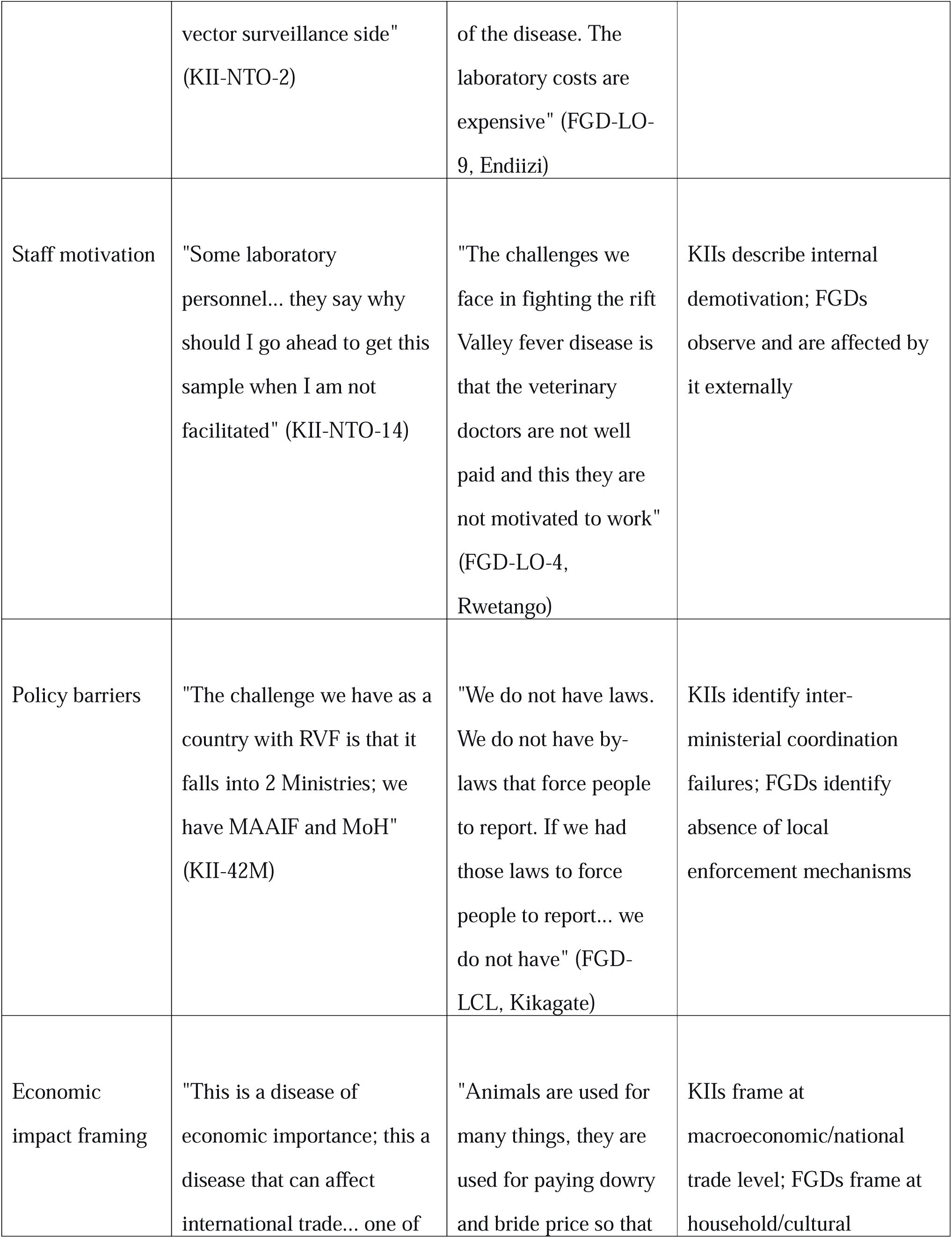
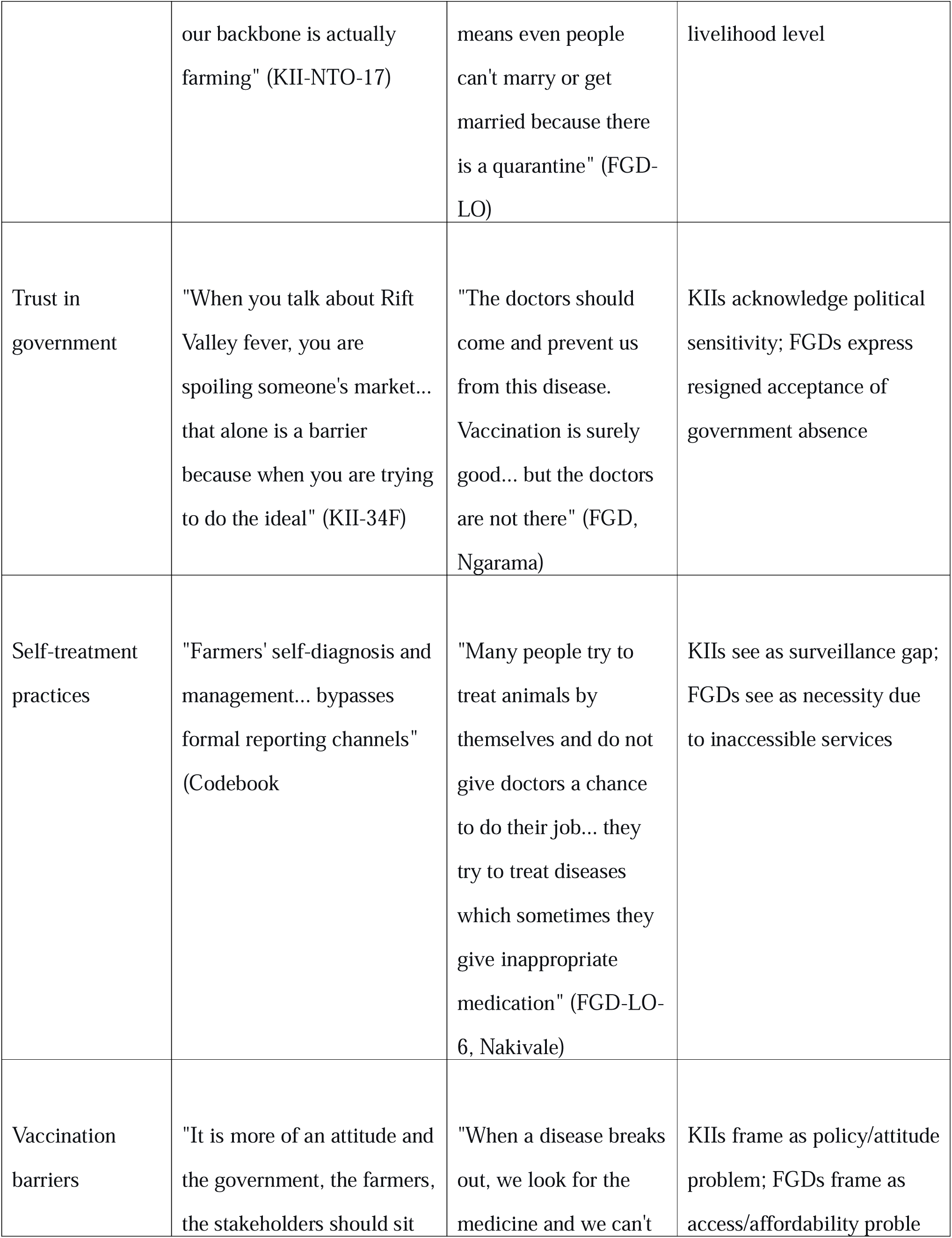
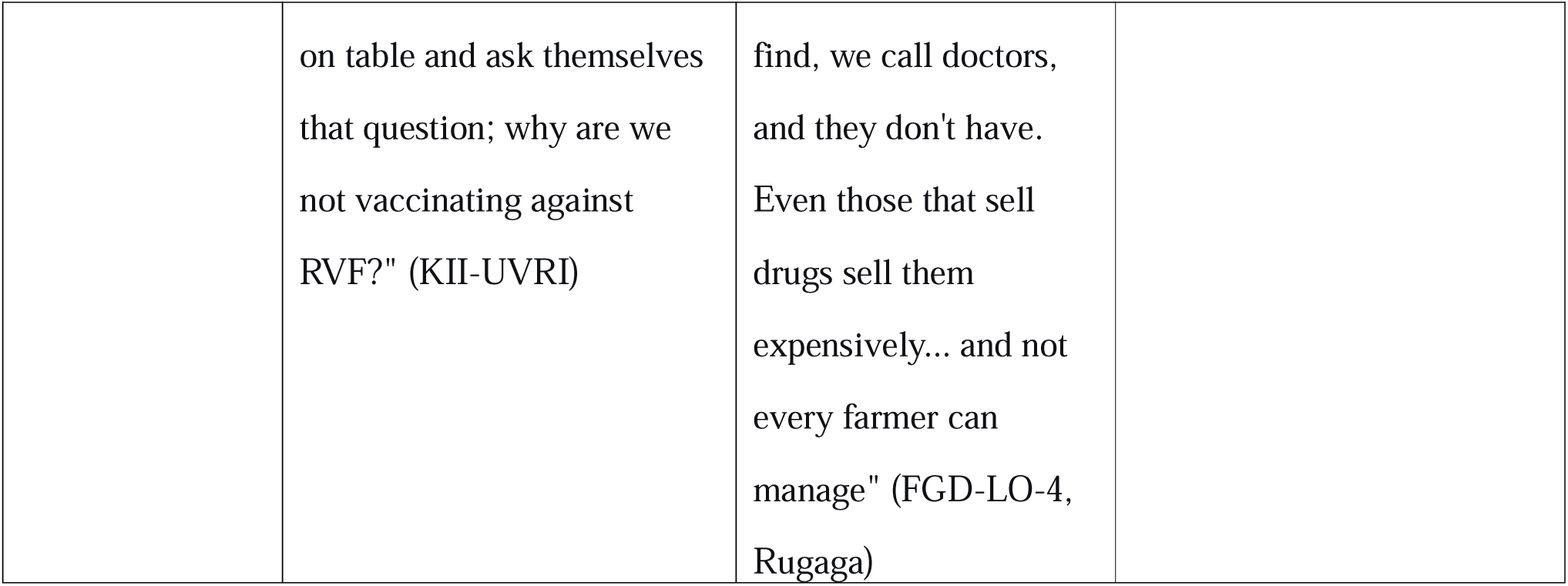

#### Unique Insights from Each Group

**Unique KII Insights (Strategic/Systemic Level):**

- Inter-ministerial coordination failures (MAAIF vs MoH)
- One Health platform weaknesses
- National laboratory capacity and hub system
- Policy inertia and political sensitivity around disease declaration
- International trade implications
- Surveillance system architecture (indicator vs event-based)
- Vaccine development and research pipeline

**Unique FGD Insights (Experiential/Community Level):**

- Daily reality of inaccessible veterinarians
- Self-medication practices as a survival strategy
- Cultural dimensions (dowry, bride price) of livestock loss
- Physical terrain and network connectivity barriers
- Distrust born from repeated reports
- Observation of demotivated staff
- Confusion between RVF and endemic diseases in practice
- Economic desperation leading to high-risk behaviours

### S6 Appendix: Ranking of Facilitators and Barriers

**Ranking of Barriers (Highest to Lowest Priority)**
| **Rank** | **Barrier** | **COM-B Domain** | **Quotes/Files** | **References** | **Justification** |
| --- | --- | --- | --- | --- | --- |
| 1 | Poor knowledge about RVF among communities | Psychological Capability | 22 | 55 | Most cited barrier (55 references) . KIIs confirm: "all confirmed RVF cases submitted as Ebola suspects." FGDs: "We don't know the disease." Foundational barrier—without recognition, no reporting occurs |
| 2 | Few staff, big district (understaffing) | Physical Capability | 8 | 18 | Severe human resource constraint. One vet for 4+ sub-counties. FGDs: "5 months without seeing the doctor." KIIs confirm no replacements for retired staff. Directly affects response capacity. |
| 3 | Absence of feedback mechanisms | Reflective Motivation | 3 | 12 | Most demotivating factor despite moderate references. KIIs: "when someone reports, they<br>should get action and feedback... most technical people have been demotivated."<br>FGDs repeatedly: "we report, but we never hear what happened."<br>Creates vicious cycle of disengagement. |
| 4 | Limited transportation means | Physical Opportunity | 8 | 13 | Surveillance staff lack vehicles; shared vehicles recalled mid-mission. KII: "this specific vehicle may be needed for other surveillance activities."<br>Directly impedes outbreak investigation |
| 5 | Confusion of RVF with other diseases | Psychological Capability | 10 | 14 | "All RVF cases confirmed were submitted as Ebola suspects." FGDs confuse with brucellosis, malaria.<br>Absence of local name compounds problem. |
| 6 | Economic sensitivity/fear of quarantine | Reflective Motivation | 4 | 5 | "When you talk about RVF, you spoil someone's market." Powerful disincentive that overrides civic duty. Farmers sell sick animals quietly rather than report. |
| 7 | Limited laboratory capacity at district level | Physical Capability | 6 | 7 | Samples must travel to UVRI/NADDEC. Delays of days to weeks. No local diagnostic confirmation capacity. |
| 8 | Poor One Health coordination | Social Opportunity | 4 | 7 | KIIs: "the most challenging part where we have not strengthened is the 'one health approach'." Human and animal health sectors operate in silos. |
| 9 | No local name for RVF | Psychological Capability | 1 | 1 | Single reference but profound implication. |
|  |  |  |  |  | KII: "community cannot label it... unless the community understands, they cannot change."<br>Prevents disease entering community discourse. |
| 10 | Farmers' self-diagnosis and management | Reflective Motivation | 5 | 7 | Bypasses formal channels. FGDs: "people were used to buying drugs from shops." Perceived as faster than waiting for unresponsive system. |
| 11 | Rough terrain and geographic remoteness | Physical Opportunity | 6 | 9 | FGDs: "when you call a doctor, he overcharges you because he is coming from very far." Roads impassable in rain; isolates communities. |
| 12 | Weak policies and regulations | Psychological Capability | 7 | 8 | FGDs: "we do not have by-laws that force people to report." |
|  |  |  |  |  | Unregulated animal movements across porous borders. |
| 13 | Delays in reporting | Physical Opportunity | 3 | 6 | FGDs: "a farmer says let me first wait and see." Passive surveillance mindset; reactive rather than proactive. |
| 14 | Limited storage capacity (cold chain) | Physical Opportunity | 3 | 4 | Vaccines and samples require cold chain; fridges inadequate or shared with human health programs. |
| 15 | Low perceived risk during inter-epidemic periods | Reflective Motivation | 0 | 0 | Mentioned in discussion; complacency sets in when no active outbreak. |
| 16 | Distrust in government response | Reflective Motivation | 0 | 0 | Implicit throughout; explicit in discussion but not coded separately. |
|  | **Ranking of Facilitators (Highest to Lowest Priority)** |  |  |  |  |
| 1 | Presence of trained staff (VHTs, CAHWs, district teams) | Physical Capability | 13 | 19 | Most cited facilitator. KIIs: trained 70+ IPC focal persons, 30+ lab officers. FGDs acknowledge accessible vets as motivator. Foundation of surveillance capacity. |
| 2 | Mobile phones and communication platforms (WhatsApp, radio) | Physical Capability | 11 | 13 | FGDs: "We have phones and we can call the vet." KIIs: district health WhatsApp groups. Ubiquitous technology enables reporting without new infrastructure. |
| 3 | Community structures (LC1 meetings, VHTs, local leaders) | Social Opportunity | 5 | 9 | FGDs: "we meet every month and discuss... we meet there with the veterinarians and report." Existing governance mechanisms provide reporting pathways. |
| 4 | Knowledge about RVF<br>(from training/experience) | Psychological<br>Capability | 20 | 29 | Trained farmers recognize signs. KIIs: "farmers who had received training were better able to identify abortions as a potential sign of RVF."<br>Modifiable capability. |
| 5 | Presence of national reference laboratories<br>(UVRI, CPHL, NADDEC) | Physical<br>Opportunity | 5 | 8 | Confirmatory diagnosis capability exists at national level. Turn-around time ~24 hours once samples arrive. Critical for outbreak confirmation |
| 6 | Fear of economic and cultural losses | Automatic<br>Motivation | 5 | 8 | FGDs: "significant economic losses due to death and abortion."<br>Livestock for dowry creates powerful intrinsic motivation to seek help. |
| 7 | Emergency Operations Centre (EOC) capacity | Physical Capability | 1 | 1 | KII: "when we receive an alert in the<br>emergency operations centre, then we verify the information."<br>Coordination hub for response. |
| 8 | Experience from past outbreaks | Reflective Motivation | 6 | 10 | Communities remember severity. KII: "preparedness Uganda... has done so well in generally outbreaks."<br>Collective memory motivates reporting. |
| 9 | Decentralized surveillance system | Physical Opportunity | 3 | 3 | Focal persons at district, HSD, facility levels.<br>KII: "right from the health facility; we have facility surveillance focal persons."<br>Structure exists despite resource gaps. |
| 10 | One Health coordination (where functioning) | Social Opportunity | 7 | 8 | KII: "we supported joint risk assessment in 2019, and country prioritized RVF." Policy-level commitment exists; |
|  |  |  |  |  | implementation at district level is weaker. |
| 11 | Presence of cold chain system | Physical Opportunity | 5 | 7 | KII: "we have fridges though they are not enough." Basic infrastructure for sample preservation and potential vaccination exists. |
| 12 | M-Trac platform and electronic reporting | Physical Opportunity | 1 | 2 | Mobile-based reporting system for human health; potential for integration. KII: "officers can now send in immediate mobile reports." |
| 13 | RVF as priority zoonotic disease | Social Opportunity | 3 | 3 | Policy recognition attracts attention and potential resources. KII: "the country prioritized RVF among the three diseases. |
| 14 | NGO support (FAO, Red Cross, medical teams) | Social Opportunity | 4 | 6 | Fill gaps where government resources insufficient. Training, transport, surveillance support. |
| 15 | Diagnostic innovation capacity | Physical Capability | 1 | 1 | KII: "we are developing an open platform where we are able to design a diagnostic using PCR."<br>Local research capacity for future solutions. |

## Notes

### Competing Interest Statement

The authors have declared no competing interest.

### Author Declarations

Ethical approval for this study was sought and received from Makerere University School of Public Health (Reference No: SPH-2022-364). Administrative clearance from the Isingiro District local government's administration was also required and provided by the Chief Administrative Officer's office (dated 20th March 2023). All methods were performed in accordance with relevant national and local guidelines and regulations. Respondents were taken through a consenting process and were provided with information about the study. Their voluntary participation and consent was recorded in writing. To enhance understanding, we translated all the data collection tools and informed consent forms into the local language (Runyankole), which was the most spoken language in the Isingiro District. All the data were de-identified to protect the identity of our informants and stored in an encrypted and password-protected external drive of the corresponding author. The emerging quotes were anonymised by using aggregated descriptors.

### Summary of Updates

We added in the comparative analysis of the dataset to show how the themes converge or diverge between the FGDs and KIIs. I also quantified the number of times the themes emerged or was mentioned.

